# Common trajectories of highly effective CD19-specific CAR T cells identified by endogenous T cell receptor lineages

**DOI:** 10.1101/2021.12.05.21266287

**Authors:** Taylor L. Wilson, Hyunjin Kim, Ching-Heng Chou, Deanna Langfitt, E. Kaitlynn Allen, Jean-Yves Métais, Mikhail V. Pogorelyy, Pratibha Kottapalli, Sanchit Trivedi, Scott R. Olsen, Timothy Lockey, Catherine Willis, Michael M. Meagher, Brandon M. Triplett, Aimee C. Talleur, Stephen Gottschalk, Jeremy Chase Crawford, Paul G. Thomas

## Abstract

Current chimeric antigen receptor-modified (CAR) T cell therapy products are evaluated in bulk, without assessment of the possible heterogeneity in effector potential between cells. Conceivably, only a subset of the pre-infusion product differentiates into optimal effectors. We generated a comprehensive single-cell gene expression and T cell receptor (TCR) sequencing dataset using both pre- and post-infusion CD19-CAR T cells from peripheral blood and bone marrow of pediatric patients with B cell acute lymphoblastic leukemia (B-ALL). We identified potent effector post-infusion cells with identical TCRs to a subset of pre-infusion CAR T cells. Effector precursor CAR T cells exhibited a unique transcriptional profile compared to other pre-infusion cells, and the number of effector precursor cells infused correlated with peak CAR T cell expansion. Additionally, we identified an unexpected cell surface phenotype (TIGIT+, CD62L^lo^, CD27-), conventionally associated with inhibiting effective T cell responses, that we used to successfully enrich for subsequent effector potential. Collectively, these results demonstrate that highly diverse effector potentials are present among cells in pre-infusion cell products, which can be exploited for diagnostic and therapeutic applications. Furthermore, we provide an integrative experimental and analytical framework for elucidating the biological mechanisms underlying effector development in other CAR T cell therapy products.

## Introduction

Chimeric antigen receptor-modified (CAR) T cells have improved treatment outcomes for many cancer types, primarily hematological malignancies. For B cell acute lymphoblastic leukemia (B-ALL) in particular, CD19-CAR T cells have exhibited remarkable activity, leading to their FDA approval in 2017. However, only a subset of patients achieves long-term remissions without subsequent consolidative allogeneic hematopoietic cell transplantation (1–4). Treatment failure is most likely multifactorial, including lack of cellular expansion post infusion, diminished effector differentiation, and variable long-term persistence to adequately control B-ALL lymphoblast proliferation. These factors are likely, at least in part, due to intrinsic features of CAR T cells themselves.

Studies have begun to elucidate the signatures within CAR T cell products that are associated with their potency. For instance, the presence of inhibitory exhaustion-associated markers on the cell surface, such as LAG3 and TIM3 on pre-infusion CAR T cells, has been associated with poor outcomes (5). Bulk RNA sequencing of apheresed T cells prior to CAR T cell production has shown that chronic interferon signaling contributes to decreased CAR T cell persistence, whereas TCF7 expression was associated with maintenance of a naïve T cell state prior to infusion and persistence of CAR T cells after infusion (6). Additionally, bulk transcriptional profiling of the infusion product showed that sustained remissions were associated with high expression of T cell memory genes in the infusion product, whereas markers of exhaustion and apoptosis were features of infusion products among non-responders (7). These and other studies have been instrumental for characterizing broad attributes of T cells and CAR infusion products that correlate with CAR T cell activity. However, understanding how specific phenotypes within heterogeneous CAR products translate to distinct functional subsets post infusion is critical for understanding how and why certain CAR T cell clones expand, persist, and become effectors within patients.

A recent study explored this issue by comparing the single-cell gene expression signatures of pre-infusion CAR T cells that proliferated to those that failed to expand once infused (8). Intriguingly, their analysis of clonal lineages from two patients with non-Hodgkin’s lymphoma (NHL) revealed that CAR T cell clones whose relative frequencies increased after infusion tended to exhibit higher cytotoxic profiles. This study demonstrated that detailed and integrative analyses spanning both pre-infusion products and post-infusion samples stand to uncover unique cellular signatures associated with optimal CAR T cell performance. Along those lines, we reasoned that identification of additional genes that poise CAR T cells to differentiate into ideal effectors, especially those that are expressed on the cell surface, could provide opportunities to enhance the efficacy of CAR T cell products.

In this study, we sampled CD19-CAR T cells in the pre-infusion product and at several time points following infusion in pediatric patients receiving treatment for B-ALL. By characterizing over 180,000 CAR T cells from infusion products (14 patients) and post-infusion samples (13 patients), we were able to dissect CAR T cell heterogeneity pre- and post-infusion and reconstruct clonal lineages across time. Leveraging an integrative approach that combines patient-focused single-cell genomics with T cell receptor sequencing, we identified and validated a unique gene expression signature present in a subset of pre-infusion product cells that gave rise to highly effective post-infusion CAR T cell phenotypes. The number of these potent effectors delivered to patients was found to correlate with the degree of CAR expansion after infusion. The shared phenotypes we characterize across patients can be used to understand the mechanisms underlying CAR T cell efficacy and to improve product generation and validation during treatment.

## Results

### Pre- and post-infusion CAR T cells have distinct gene expression profiles

To determine features of effective CAR T cell therapies, we undertook a comprehensive gene expression profiling of 16 pediatric patients with relapsed/refractory B-ALL, 15 of whom had received autologous T cells expressing a CD19.4-1BBz CAR (CD19-CAR) (9, 10) post lymphodepleting chemotherapy on our investigator-initiated clinical study (NCT03573700; **Fig 1A**). Patients 0 through 12 have been reported in detail elsewhere (11). Twelve out of the 15 patients who were infused with CAR T cells achieved a complete response (CR), with 11/12 CRs being minimal residual negative (MRDneg) at four weeks post CD19-CAR T cell infusion (see **Supplementary Clinical Information**). We analyzed the CD19-CAR T cell product generated in our good manufacturing practice facility (GMP product), as well as post-infusion samples obtained from peripheral blood mononuclear cells (PBMCs) and bone marrow aspirates at regular intervals. Single-cell gene expression and T cell receptor (TCR) sequencing were performed on sorted T cell populations in the GMP product and at weeks 1-4, 8, and months 3 and 6 post infusion. Across all patients and time points, we sequenced 118,749 GMP product CAR T cells and 66,042 post-infusion CAR T cells, with an average of 11,549 cells per patient (SD = 7,335) and 20,532 cells per time point (SD = 37,898). Of note, the month 6 post infusion time point only consists of 7 CAR T cells, all derived from a single patient (**Supplementary Table 1**). GMP product CAR T cells represented 64% of the total CAR T cell population and exhibited on average 3,320 genes per cell (SD = 856). Post-infusion CAR T cells were less transcriptionally active, expressing 2,299 genes per cell on average (SD = 931).

**Figure 1.**
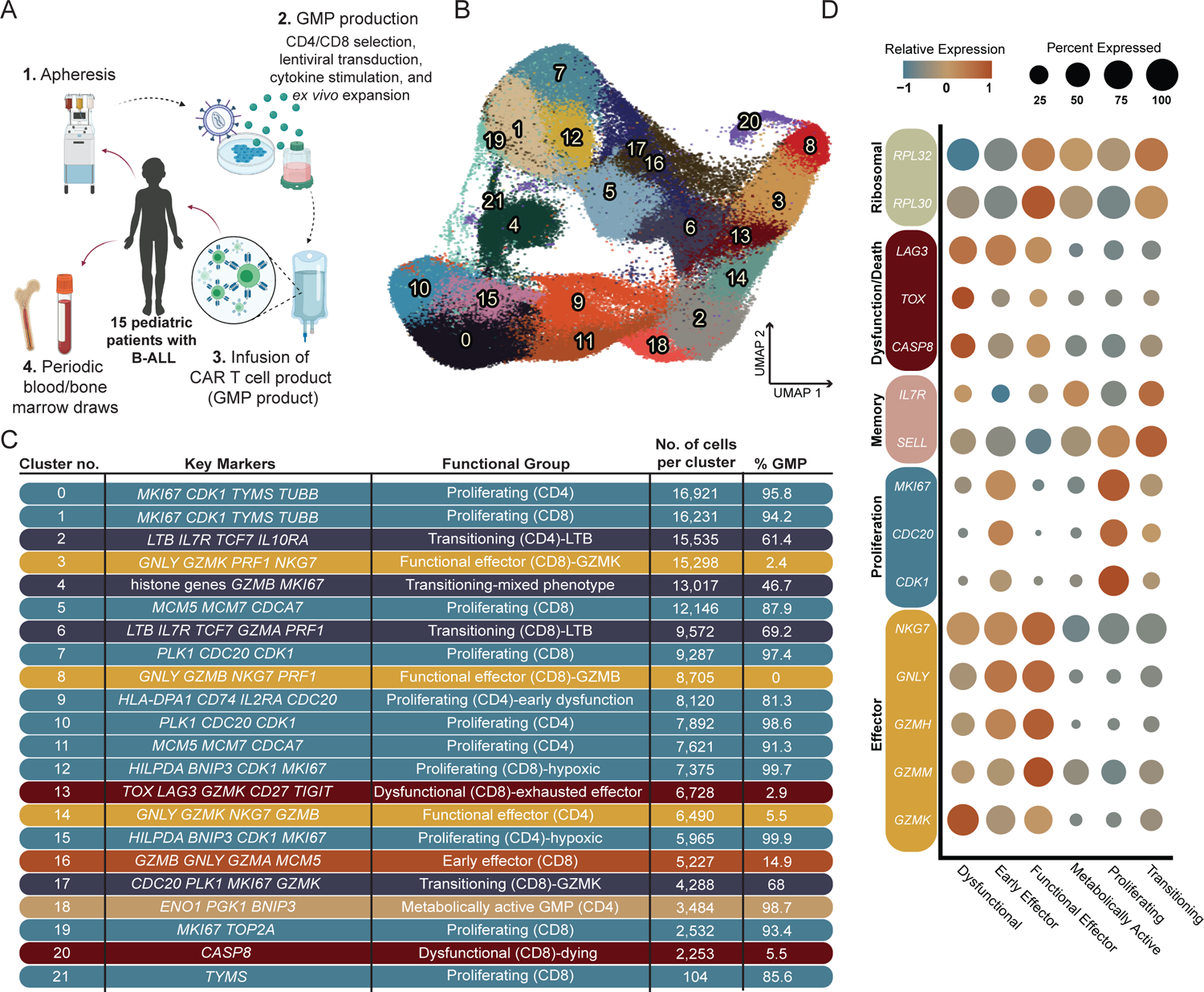
Identification of transcriptional subsets within pre- and post-infusion CAR cells. **A**, Schematic of clinical trial process. Leukocytes were apheresed from 16 pediatric patients undergoing CD19-CAR T cell therapy. T cells were selected, virally transduced with the CAR-containing lentivirus, and expanded. The autologous CAR T cell products were infused into 15 of the patients, and blood and bone marrow were drawn at protocol-specific time points to isolate CAR T cells. **B,** UMAP plot with shared nearest neighbor clustering of 184,791 pre- and post-infusion CAR T cells across all patients, colored by 21 transcriptional clusters. **C,** Chart depicting the cluster number as indicated in panel A, with key genes used to characterize the transcriptional profile of each cluster, the functional groups each cluster was assigned to based on the specific transcriptional profile, the number of cells in each cluster, and the percent GMP composition of each cluster. Colored bars correspond to each broad functional group (proliferating: slate blue; transitioning: navy blue; early effector: burnt orange; functional effector: dark yellow; dysfunctional: burgundy; metabolically active: light brown). **D**, Dot plots showing the relative expression of genes characteristic to relevant cellular processes as listed. Dot size corresponds to the percent of cells expressing each gene.

To understand the range of CAR T cell phenotypes and how these phenotypes change from the pre-infusion product throughout the course of treatment, we performed UMAP dimensionality reduction and shared nearest neighbor clustering based on gene expression profiles (**Fig 1B**). Included in this broad analysis were both GMP product and post-infusion CAR T cells, which were generally segregated across the primary axis of variation **(Fig S1A)**. Unsurprisingly, CD4+ and CD8+ CAR T cells also separated distinctly, with few clusters shared by both T cell types **(Fig S1B)**. Differential gene expression analysis across the clusters revealed several distinct T cell functional states. Clusters predominated by GMP product CAR T cells were classified as proliferative with high expression of genes such as *MKI67*, cell-cycling genes *CDK1* and *CDC20*, DNA replication genes *MCM7, TOP2A*, and *TYMS* (CD8: clusters 1, 5, 7, 12, 19, 21; CD4: clusters 0, 9, 10, 11, 15; **Fig 1C, D**). In contrast, a subset with relatively lower proliferation signals but high expression of *ENO1, PGK1*, and *BNIP3* was identified as metabolically active CD4+ GMP product cells (cluster 18; **Fig 1D**).

Within post-infusion CAR T cells, we observed several functional effector T cell populations characterized by robust expression of cytotoxic genes *GNLY, PRF1*, and *NKG7* (CD8: clusters 3, 8, and 16; CD4: cluster 14; **Fig 1B-D**). Interestingly, expression of *GZMK* distinguished cluster 3 from cluster 8, the classic effector *GZMB*-expressing CD8 effector population (**Fig S1C**). Others have previously suggested that *GZMK*-expressing T cells are a distinct T cell population (12), and we found that this cluster was more heterogeneous in expression space compared to the tightly grouped cells from cluster 8 (**Fig 2A; Fig S1D**). A separate, early functional effector cluster (16) was distinguished by upregulation of *MCM5*, indicating a maintenance of the proliferation signals seen in GMP product CAR T cells despite their simultaneously upregulated effector genes (**Fig S1E**). We also identified two dysfunctional CAR T cell post-infusion populations unique to CD8s, clusters 13 and 20. High expression of a suite of inhibitory markers within cluster 13, including *TOX, LAG3*, and *TIGIT*, suggested T cell exhaustion (**Fig 1B-D**). In contrast, cluster 20 CAR T cells exhibited apoptotic processes, indicated by downregulation of genes needed in translation, ribosomal subunit genes *RLP30* and *RPL32*, and high expression of *CASP8* (**Fig 1C**). Despite the presence of substantial clusters characterized by proliferating CD4+ CAR T cells within the GMP product, CD4+ CAR T cells made up only 25.7% of post-infusion cells profiled. Similarly, the size of the CD4 effector compartment was much smaller than that of the CD8 CAR T cells post infusion, likely stemming at least in part from the presence of a proliferating CD4 cluster (9) characterized by early dysfunction.

**Figure 2.**
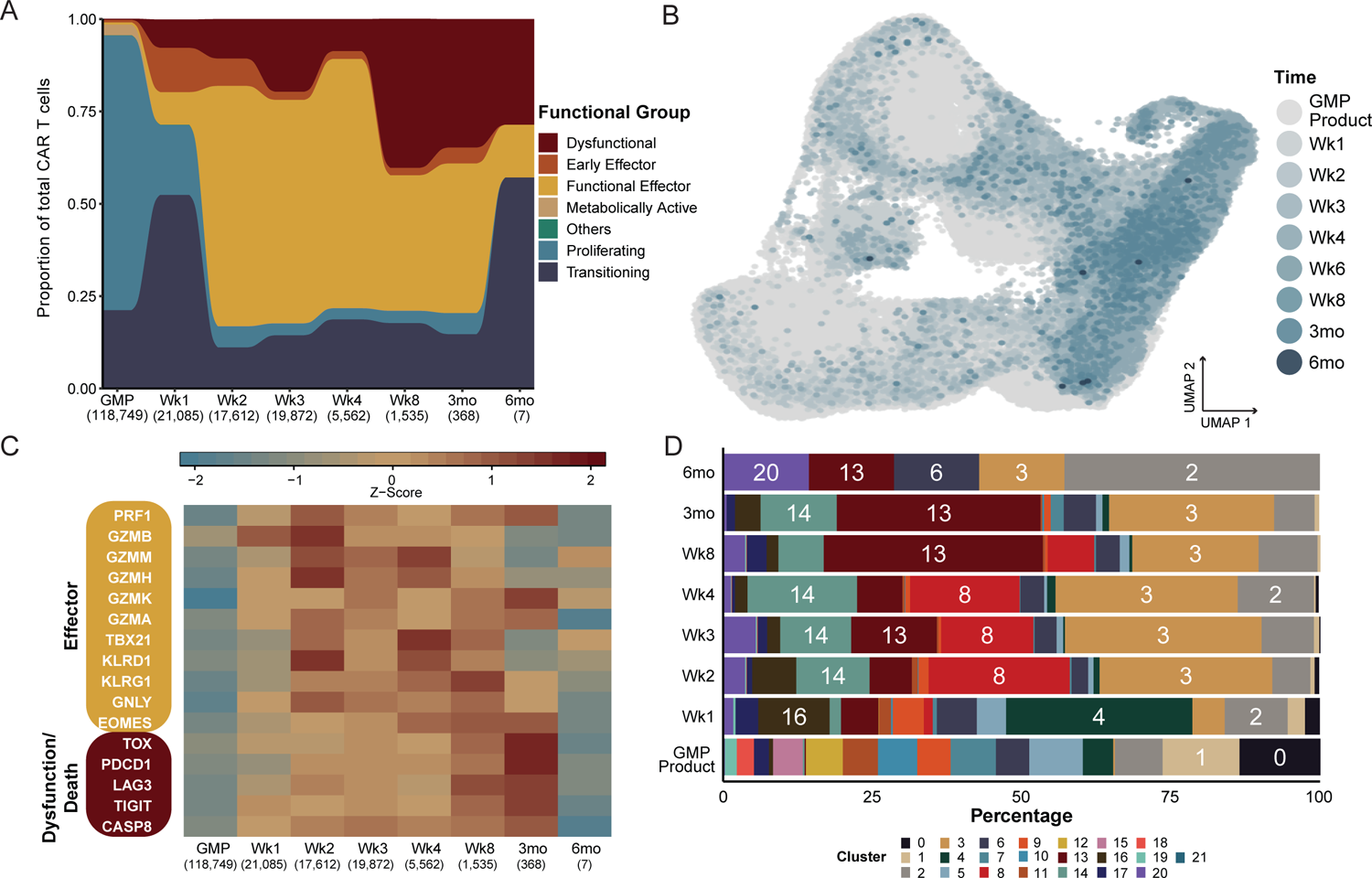
Expression of effector and dysfunctional genes over time correlates with kinetics of CAR T cell subsets. **A**, Relative proportion of functional groups as defined in Figure 1, aggregated across donors for each pre-infusion and post-infusion time point. Number of cells per time point are included in parentheses under each time point label. **B**, UMAP of pre- and post-infusion CAR T cells, colored by GMP status or post-infusion time point. Cells from later time points were plotted on top of those from earlier time points. **C**, Heatmap of average gene expression across CAR T cell time points, visualizing variation in genes associated with cytotoxic effector function or T cell dysfunction as indicated. **D**, Stacked bar plots portraying each transcriptional cluster’s relative contribution to the indicated time points, colored by transcriptional cluster.

Although most transcriptional clusters were strongly associated with either pre- or post-infusion samples, some populations (CD8: 6 and 17; CD4: 2; **Fig S1E**) instead exhibited a putative transitional state between the proliferative signature associated with GMP product CAR T cells and the cytotoxic effector differentiated state of the post-infusion CAR T cells. These transitional phenotypes were identified by the joint expression of several cytotoxic genes, including *LTB* (clusters 2 and 6), *GZMA* (cluster 6), *GZMK* (cluster 17), and *PRF1* (cluster 6), in addition to the expression of proliferation signals *CDC20*, *PLK1*, and *MKI67* (**Fig S1E**). Lastly, we identified a putative hybrid population, cluster 4, that contained both GMP product and post-infusion cells with mixed CD4+ and CD8+ T cell subsets.

Overall, assessments of variation in gene expression allowed us to identify distinct transcriptional programs between GMP product and post-infusion CAR T cells. Signals associated with initiation and maintenance of the complex pathways involved in T cell proliferation were evident within both CD4+ and CD8+ GMP product CAR T cells, while post-infusion CAR T cells demonstrated signs of differentiation into effector T cells displaying potent cytotoxic transcriptional profiles.

### Post-infusion CAR T cells become more effector-like over time

Having established that GMP product and post-infusion CAR T cells encompass several distinct transcriptional subsets, we next determined the proportion of post-infusion CAR T cells that fit within these broad transcriptional categories. Starting at week 2 post infusion, the majority of CAR T cells could be attributed to functional effector subsets (CD8: clusters 3 and 8; CD4: cluster 14; **Fig 2A**). Notably, the week 2 sample also coincided with the time of peak expansion in 8 of the 12 responding patients with available qPCR assays of CAR abundance (**Supplementary Table 2**). Because cytotoxic effectors predominated the post-infusion CAR T cell functional groups, we sought to characterize the kinetics of CAR T effector differentiation by assessing variation in the transcriptional signatures across post-infusion time points. To do this, we considered post-infusion cells based on their sampling time point but visualized in UMAP space. As expected, cells obtained from earlier time points tended to cluster near cells from the GMP products, whereas cells from later time points were more enriched near the bulk of the post-infusion populations (**Fig 2B**). Next, we characterized the development of the effector signature across GMP products and post-infusion time points. Compared to CAR T cells in the GMP products and at week 1 post infusion, genes associated with the functional effector signature were most highly expressed starting at week 2 (**Fig 2C**). This correlates with a dramatic relative expansion of the populations we identified as functional effectors (CD8: 3 and 8; CD4:14) at week 2, as they represented a total of 65% of week 2 CAR T cells (**Fig 2D**). Interestingly, the CD8+ *GZMK* expressing functional effector population (cluster 3) was maintained throughout the remainder of the sampling window and constituted a sizable 27.7% of CAR+ cells at month 3 post infusion. In contrast, the *GZMB* expressing effector group (cluster 8) was lost after week 8, concurrent with the loss of *GZMB* expression at week 8 across all clusters (**Fig 2C**). Despite CD4+ CAR T cells being the minority in the post-infusion CAR T cell compartment, the proportion of CD4+ cytotoxic effectors (cluster 14) was fairly consistent throughout the study period and encompassed 12.8% of all CAR T cells at month 3 after infusion (**Fig 2D**).

Dysfunctional T cell responses, including exhaustion, can arise in some cases of prolonged activation and effector function (13). We therefore investigated whether markers of T cell exhaustion and cellular death, identified within dysfunctional clusters 13 and 20, appeared at a time congruent with extended effector function. As early as week 1, markers often associated with exhaustion and apoptosis were observed in post-infusion CAR T cells; however, later time points (week 8 and month 3) were substantially more enriched for exhaustion and apoptotic genes (**Fig 2C**). This is supported by the time points in which the exhausted CD8+ T cell cluster (cluster 13) appears after infusion, as it constituted only a minor proportion of the population at week 1 (6.3%), increased to 14.4% at week 3, and made up 36.8% and 34.2% at week 8 and month 3, respectively (**Fig 2D**). *GZMK* was also expressed highly at month 3 post infusion, concordant with an increase in exhaustion signals; notably, this signature has been associated in other studies with precursor exhausted T cells (Tpex) (14). The dying population (cluster 20) is especially enriched at week 3 and month 6 post infusion, yet this phenotype is also present in earlier post-infusion time points (**Fig 2D**). Although the enrichment of dysfunctional signals later suggests that a subset of dysfunctional CAR T cells arise as a result of chronic effector function, evidence of dysfunction appearing as early as week 1 post infusion may indicate that some cells develop their dysfunctional state almost immediately after infusion.

### Pseudotime identifies two distinct trajectories for post-infusion CAR T cell differentiation

Comparing gene expression clusters to sample kinetics demonstrated a general trend towards effector differentiation, which accelerated at 2 weeks after infusion. However, these data also revealed surprising heterogeneity in gene expression profiles, with dysfunctional cells present even at the very earliest time points after infusion. To explore the relationship between cell fates and differentiation states, in particular the precursors and offspring of potent functional effectors (CD4: clusters 14; CD8: clusters 3 and 8) and dysfunctional cells (CD8: clusters 13 and 20), we utilized pseudotime analysis (on a downsampled subset of the data; see methods), where each cell is assigned a putative degree of progression along an inferred trajectory. Cells that fall within a specific area of the pseudotime are grouped into states.

CAR T cells from GMP products and post-infusion samples clustered along 7 distinct states (**Fig 3A**), with the root of the trajectory in state A (**Fig S2A**). Unsurprisingly, the majority of GMP product CAR T cells fell within the root of the pseudotime trajectory, while post-infusion cells predominated among the branching trajectories (**Fig 3B**). As cells progress along the pseudotime trajectory, one group (state B) splits into a distinct differentiation pathway from the others. Because state B arises earlier in pseudotime than the other states, we classified state B as “early” and states D, E, F, and G as “late.” Of note, dysfunctional cluster 13 spanned both the early and late transcriptional states, whereas dysfunctional cluster 20 was primarily confined to state B (**Fig S2B**). Due to the association of cluster 20 with cellular death processes, we evaluated the expression of the apoptotic gene *CASP8* across all states. Comparatively, expression of *CASP8* was highest in state B relative to all other states (**Fig 3C**). Thus, the pseudotime analysis indicated that a subset of cells after infusion directly acquire a dysfunctional phenotype and proceed to cluster 20 (state B; **Fig S2B**), which likely results in cell death. In contrast, another trajectory terminates in a potent effector phenotype (states C, D, and G; **Fig 3D**), while a subset of those proceeds on to exhaustion, as characterized by higher relative expression of *TOX* and *LAG3* (state C; **Fig 3C**). These interpretations broadly concur with our traditional gene expression analysis that indicated dysfunctional phenotypes developed in a subset of cells early after infusion.

**Figure 3.**
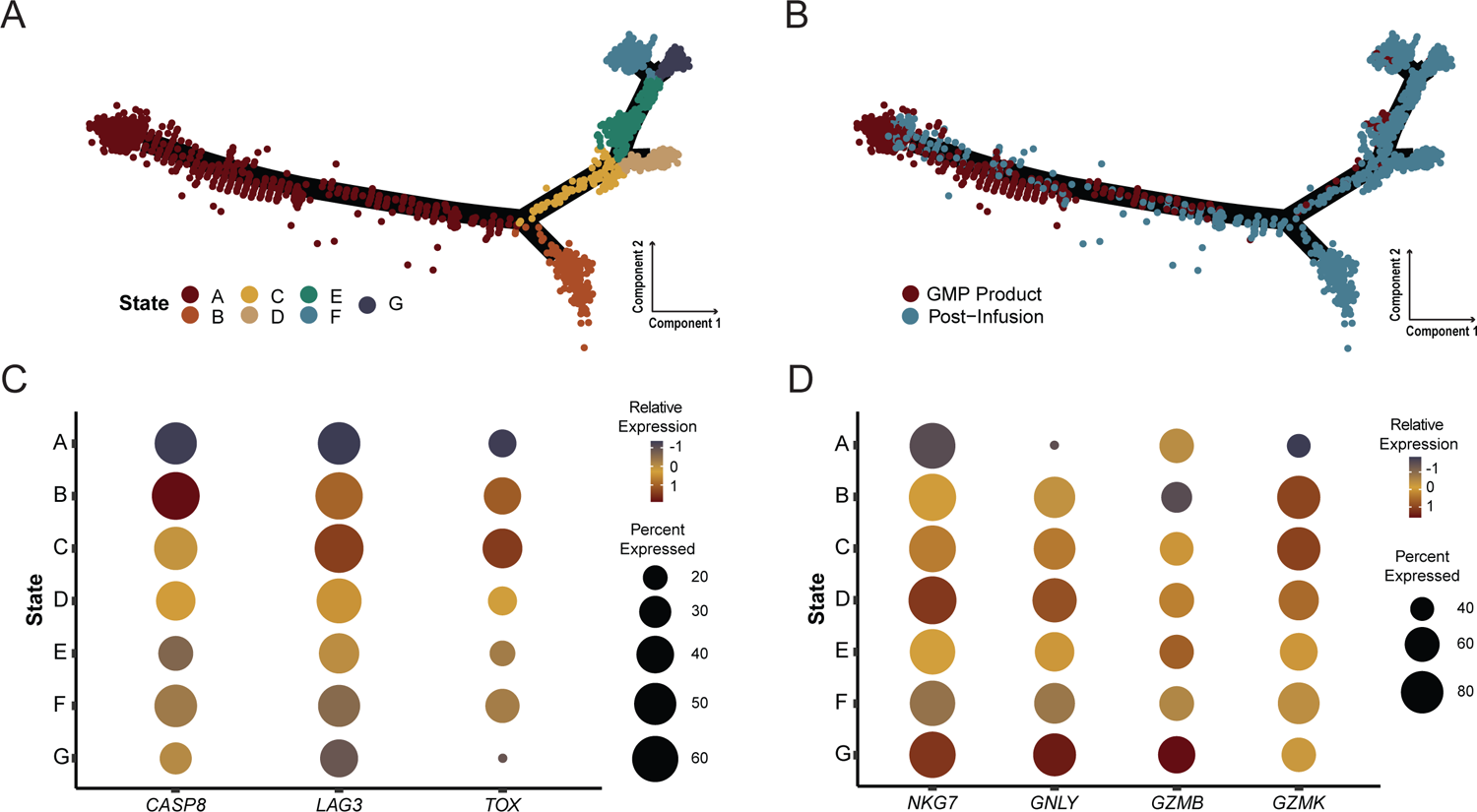
Pseudotime trajectory analysis identifies a subset of dysfunctional post-infusion CAR T cells that arise directly from the GMP product rather than from prolonged antigen exposure. **A-B**, Monocle pseudotime map depicting trajectory analysis of 3,416 CAR T cells. Downsampling was necessary due to computational limitations. Analysis included 368 cells (the number of all cells at the month 3 time point) from each time point, as well as all 840 cells with TCRs matching known pre- and post-infusion lineages regardless of cluster designation. Pseudotime states were generated based on internal clustering by the pseudotime analysis. **A**, Cells are colored by pseudotime state. **B**, Cells are colored by either GMP or post-infusion status. **C**, Dot plot comparing relative expression of *CASP8*, *LAG3*, and *TOX* across pseudotime states, with percent of cells expressing a gene encoded by dot size. **D**, Dot plot comparing relative expression of effector genes (*NKG7*, *GNLY*, *GZMB*, and *GZMK*) across pseudotime states, with percent of cells expressing a gene encoded by dot size.

### The TCR serves as a barcode for CAR T cell lineages

While pseudotime is a useful tool to infer lineage relationships and identify potential differentiation branch points, to understand what transcriptional signatures give rise to the desired effector CAR T cells, we needed a method to definitively track these lineages between pre- and post-infusion samples. The endogenous TCR is an optimal marker of T cell lineages, as it remains static throughout differentiation and is passed to each daughter cell during T cell proliferation, a process known as clonal expansion. It is estimated that most paired αβ TCRs are represented in the naive human repertoire only a small number of times, generally once (15–17). Therefore, clonal expansions, defined by cells with at least one matching α and one matching β chain (**Fig S3A**), in the effector and memory repertoires have a high likelihood of being descended from the same parental clone. Thus, we hypothesized that we could utilize the endogenous TCR in transduced cells as a lineage barcode for CAR T cells after product generation and infusion.

To test whether we could track CAR T cell clonotypes across multiple time points, we first quantified the number of unique clones in GMP products and at each post-infusion time point. Targeted cDNA enrichment of the complementarity-determining region 3 (CDR3) of both α and β subunits of the TCR yielded a total of 153,853 unique αβ pairs. Cells with a given αβTCR appearing at more than one time point were defined as “lineages.” Although the lineages identified by this classification are persistent across time points, it is important to note that these lineages are undersampled due to the fact that we cannot exhaustively sample the infused cells within a patient; importantly, in subsequent analyses this undersampling biases our results towards the null hypothesis that persistent and non-persistent lineages are similar.

In 10 out of 15 infused patients, we tracked multiple lineages originating in the GMP product with a range of 4 to 125 (**Fig 4A**; **Supplementary Table 3**). Overall, investigations into relative clonal dynamics of CAR T cells over time uncovered considerable diversity in the GMP product. After infusion, evidence of clonal expansion (clone size > 10 cells) occurred in 7 patients (**Supplementary Table 4**). We observed a range of clone sizes among CAR T cells across the post-infusion time points. For instance, 44,981 TCRs (defined using the one-from-each approach) assayed post infusion were only observed once. However, we also detected 2,620 TCRs more than once but in <= 10 cells. Analyzing these data across all patients, we observed a difference between average clone sizes in persistent clonal lineages compared to clonotypes that we only observed once (**Fig S3B**), with persistent lineages exhibiting larger overall clone sizes. This suggests that these lineages represent expanded or expanding populations.

**Figure 4.**
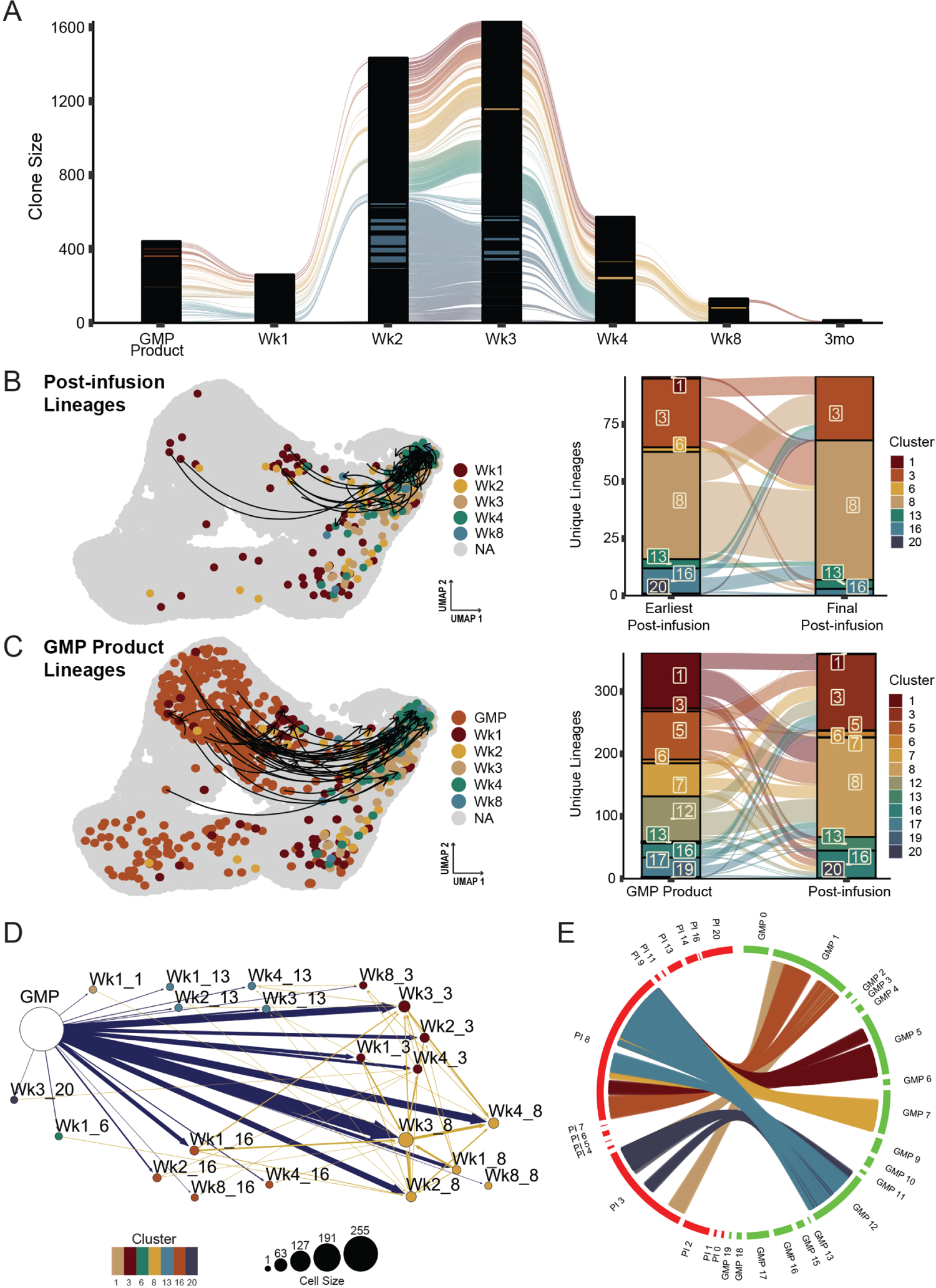
Tracking of endogenous TCR over time identifies CAR T cell lineages and their subsequent fates. **A**, Alluvial plot of CAR T cell lineages across the GMP and post-infusion (PI) time points. Lineages were defined using a “one-from-each” approach, where cells that match their most highly expressed (as a stringency filter) α and β chains are designated as lineages (detailed further in Fig S3). Each line corresponds to an individual CAR T cell lineage. Due to space constraints, only immediately consecutive connections are visualized (e.g., excluding direct connections between GMP and Wk3). Black columns result from the stacking of many clones that are detected in only a single cell at a given time point. **B** (left), UMAP plot emphasizing CAR T cell lineages detected across PI time points. Arrows indicate the CD8+ CAR T cells of the same lineage, starting at the earliest PI time point a lineage was detected and ending at the final time point a lineage was detected. Cells without lineages across PI time points are colored grey. All other cells are colored by their post-infusion time point. Colored cells without arrows are in lineages that span GMP to PI time points. **B** (right), Alluvial plot depicting the cluster assigned to the earliest post-infusion detection of a lineage and the cluster assigned to the latest detection of a lineage. Colors correspond to transcriptional clusters. When lineages span multiple clusters at the same time point, we include both clusters in the plot. **C** (left), UMAP plot emphasizing CAR T cell lineages spanning GMP and multiple post-infusion time points. Arrows indicate the CD8+ CAR T cells of the same lineage, starting at the first detection of the lineage in the GMP and ending at the final detection of the lineage. To aid in visualization, only CD8+ GMP lineages observed in more than one post-infusion time point were represented with an arrow. Cells without lineages tracking to the GMP product are colored grey. All other cells are colored according to their GMP status or post-infusion time point. **C** (right), Alluvial plot depicting the cluster assigned to the lineage in the GMP sample and the cluster assigned to the final detection of the lineage. Colors correspond to transcriptional clusters. When lineages span multiple clusters at the same time point, we include both clusters in the plot. **D**, Network plot of CAR T cell lineages. Arrows link cells that share TCRs, directed from earlier to later time points (blue: GMP to PI red: PI to PI). Dot size indicates number of cells, dot colors correspond to transcriptional clusters, and arrow width increases as the number of lineages increases. Labels indicate either GMP status or the time point followed by the transcriptional cluster number. **E**, Circos plot visualizing most frequent lineage connections between GMP clusters and PI CD8+ functional effector clusters (3 and 8). Each position on the outer ring represents a cell, and each line represents a lineage. When lineages span multiple GMP or PI clusters, all clusters are plotted. For ease of visualization, lineages between GMP-PI clusters with fewer than 50 total connections are excluded.

To determine the trajectories of particular post-infusion cell states, we first identified the earliest time point that a specific TCR lineage was observed, determined the cluster encompassing that clone at that earliest time point, and then determined the cluster that housed the final time point where that TCR clonotype was observed (**Fig 4B**). We focused on the lineages across CD8+ clusters because very few lineages were identified within the CD4+ T cells in general (and zero CD4+ lineages crossing multiple post-infusion time points), reflecting a relative underabundance of CD4+ CAR T cells observed after infusion. Interestingly, multiple αβTCRs mapped across the functional effector clusters 3 and 8, potentially suggesting that CD8+ functional effector subsets share common origins. For instance, CAR T cells within cluster 3 (*GZMK+* effectors) at relatively early time points often shared TCRs with cells from cluster 8 (*GZMB+* effectors) at later time points, again suggesting that GZMK+ expressing CD8+ T cells can be an earlier effector state immediately preceding cytotoxic effectors (18). However, we also observed evidence of the opposite phenomenon occurring, where *GZMB*-expressing cluster 8 lineages later acquire the *GZMK+* effector signature. Furthermore, pseudotime analysis suggested that dysfunctional cell states, consisting primarily of clusters 13 and 20, arise from two transcriptional pathways: early dysfunction rapidly acquired after infusion, or late dysfunction acquired as a consequence of sustained effector function. We observed some post-infusion lineages in clusters 3 and 8 at early time points that were also seen in cluster 13 at later time points, which may represent the conventional loss of effector potential and onset of exhaustion in the functional effectors, or late dysfunction. Some lineages from CD8+ GMP cells, specifically from cluster 1, were mapped to cluster 13 upon infusion (**Fig 4C**), suggesting early dysfunction. However, establishing conclusive evidence of early dysfunction using TCR lineages remains complicated by relatively limited sampling of extremely diverse populations.

We next endeavored to use TCR lineages to identify precursors within the GMP that gave rise to the predominant functional effector populations observed throughout post-infusion time points (**Fig 4C**), once again concentrating on CD8+ lineages due to the low frequency of CD4+ lineages in our dataset. We identified post-infusion clonotypes that shared exact αβ TCRs with GMP cells. Because the effector populations exhibit distinct transcriptional signatures, we hypothesized that precursor lineages within the GMP would fall within a limited set of clusters. However, these effector lineages linked to several GMP clusters, including 1, 5, 7, and 12 (**Fig 4D-E**). Therefore, based on global gene expression analysis alone, no single GMP product cluster contained the precursors for potent effector lineages.

### Effector GMP product cell precursors have an identifiable signature pre-infusion

Since global gene expression analysis did not identify a single effector precursor transcriptional cluster within the GMP product, we hypothesized that more subtle gene signatures that were obscured from broader analyses might characterize a pre-effector phenotype. To test this hypothesis, we compared the transcriptional profiles of GMP product cells that shared TCRs with cells found in post-infusion effector clusters 3 and 8 to all other CD8+ CAR T cells in the GMP product whose TCRs were never observed in those clusters (GMP controls). This comparison identified both upregulated and downregulated genes associated with the precursors that share lineages with functional effector responses (**Fig 5A**). Particularly, lineage-defined effector precursors were, prior to infusion, already expressing genes associated with an effector T cell phenotype, including classic effector markers *EOMES, GNLY, GZMH, GZMK,* and *IFNG*. Surprisingly, genes that are often associated with impaired T cell activation and effector function, such as *TIGIT* and *LAG3*, were also upregulated among the effector precursors when compared to the GMP controls. Other downregulated genes within the effector precursors relative to control cells were associated with T cell memory subsets. For example, effector precursors significantly downregulated central memory genes *SELL*, which encodes for CD62L, and *IL7R*, as well as the stem cell memory gene *LEF1*. Although previous CD19-CAR T cell studies have identified CD27 as a marker for optimal CAR T cell performance (6, 7), in this analysis the GMP product T cell clones that also appear in post-infusion effector clusters exhibited markedly lower levels of *CD27* gene expression in the product.

**Figure 5.**
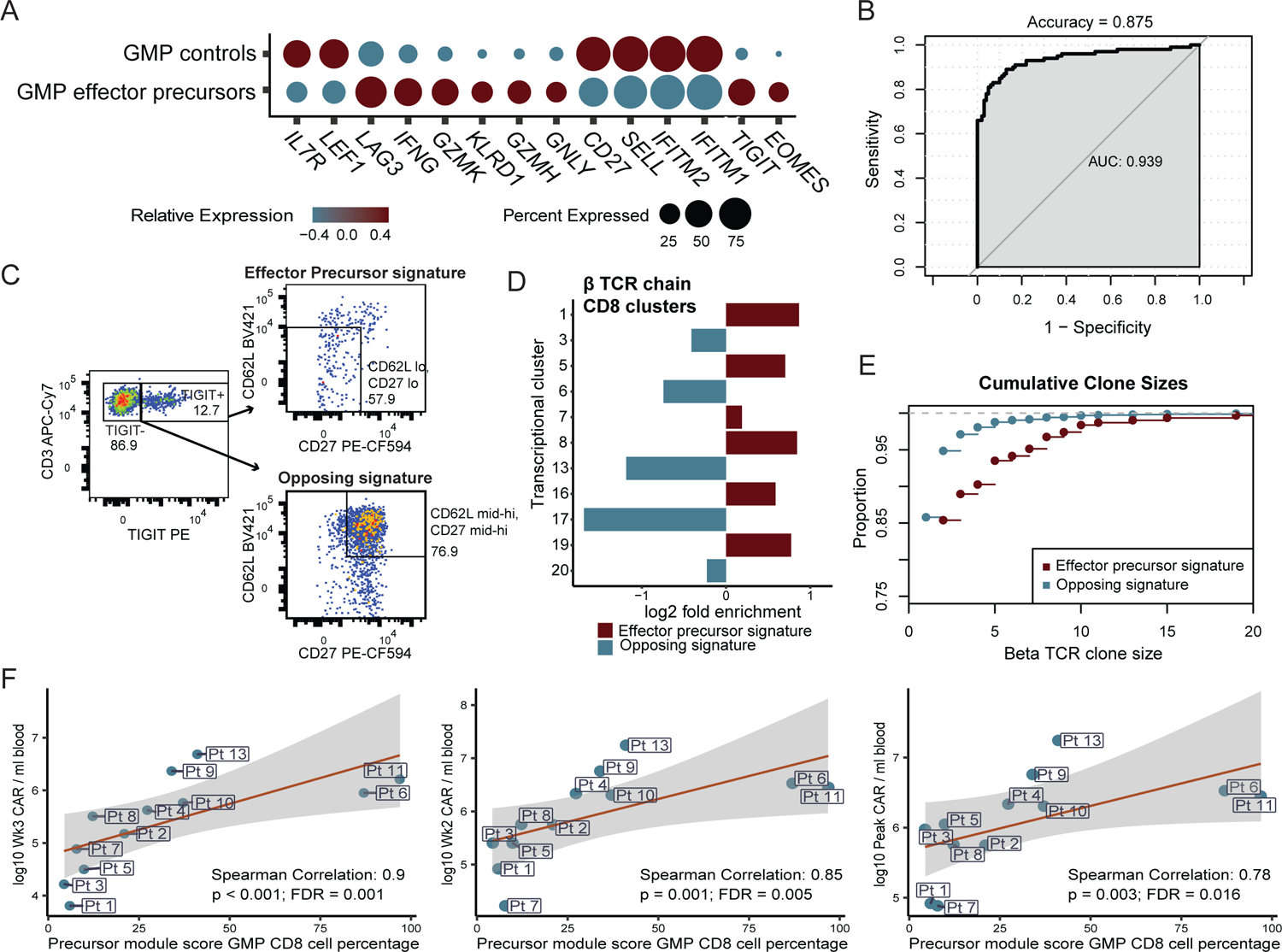
A subset of GMP CAR T cells is uniquely poised to give rise to functional effectors. **A**, Dot plot comparing relative expression of 14 genes differentially expressed between GMP effector precursors (as defined by αβTCR lineage tracing) and all other CD8+ GMP CAR T cells. **B**, AUC of the best performing iteration of an SVM classifier trained on the top 100 differentially expressed genes between these GMP effector precursors and all other CD8+ GMP T cells. The single-cell dataset was randomly downsampled with LOOCV for 1,000 iterations. **C**, Flow cytometry data from an aliquot of patient 11’s GMP infusion product, visualizing CD8+ CAR T cells with the predicted effector precursor profile (TIGIT^+^, CD62L^lo^, CD27^lo^) and the opposite non-effector associated surface profile (TIGIT^-^, CD62^hi^, CD27^hi^). **D**, Bar plot comparing the proportion of bulk β TCRs sequenced that matched those from post-infusion CD8 transcriptional clusters. Differences are represented as log_2_ fold change for the proportions of TCRs matching each cluster. Colors correspond to the precursor effector signature (red) or the opposing, non-effector signature (blue) as defined in panel C. **E**, Cumulative β clone sizes of post-infusion CAR T cells that share β TCRs with GMP product CAR T cells sorted by the precursor effector signature (red) or the opposing, non-effector signature (blue). Graph represents the proportion of cells (y-axis) cumulatively encompassed by increasing clone sizes. **F**, Correlation plots comparing the percentage of inferred effector precursors in the GMP product to CAR abundance as assayed by qPCR and represented in terms of CAR copies per ml of blood, at week 3, week 2, and peak expansion time points.

As we were able to find a profile unique to pre-effector GMP product T cells, we next tested whether this signature could be used to train a classifier that could assess CAR T cell effector potential in other GMP products. The top 100 differentially expressed genes between downsampled pre-effector and other downsampled CD8+ CAR T cells of the GMP product were selected to train a machine learning algorithm to predict whether a given CAR T cell in the GMP product would give rise to a lineage that was observed in the CD8 effector pool after infusion. LOOCV (leave-one-out cross validation) was used to validate the accuracy of the model, and the differentially expressed signatures were also computed only from each training set. Due to the immense size of the dataset, we downsampled the data for the classification analyses. Using 1,000 randomly downsampled sets of CAR T cells, the best performance of the classifier reported an accuracy of 87.5% and an AUC of 0.939, with an overall average of 78.4% accuracy and an AUC of 0.876 across the 1,000 iterations (**Fig 5B**).

We theorized that if the GMP product cell precursor signature indeed correlates with the eventual development of functional effector CAR T cells, selectively isolating these precursor cells should enrich pre-effector associated TCR clonotypes. To test this, we selected a subset of genes that encode for surface proteins that were sufficiently differentially expressed between pre-effector CAR T cells and all other CAR T cells from the GMP products (**Fig S4A**): *TIGIT*, *SELL* (CD62L), and *CD27*. CD8+ CAR T cells that contained the predicted effector precursor profile (TIGIT+, CD62Llo and CD27neg) were sorted from a cryopreserved GMP sample from a single patient. TIGIT+ cells represented 12.7% of CD8+ CAR T cells, and we further subsetted on cells that were low for both CD62L and CD27 (57.9% of TIGIT+ cells). We also sorted on CD62L and CD27 mid-hi expressing TIGIT-cells (74.9% of TIGIT-cells) to use as a population representing an opposing, non-effector precursor phenotype (**Fig 5C**). We next sequenced the complementarity determining region 3 (CDR3) of the β TCR chain within the bulk RNA obtained from each sorted population to compare the relative abundance of effector precursor lineages in each sorting scheme. Because of dramatic differences in sample sizes and the extreme diversity of the repertoires, we first compared the 1,000 most frequently observed β TCRs from each sort condition to those observed in our single-cell post-infusion data; interestingly, the top clones from the effector precursor sort were more than twice as likely to be observed post infusion than the top clones from the non-effector sort (effector: 132; non-effector: 59; Fisher’s exact test, p < 0.001). When considering all TCRβ nucleotide sequences from bulk-sorted T cells that matched those in the patient’s post-infusion CD8 repertoire, we observed marked differences in the proportion of the inferred transcriptional clusters to which each sort signature mapped (**Fig S4B**). The precursor effector signature-sorted cells expressed TCRs that were more likely to be observed in proliferating CD8 clusters (1, 5, 7, 19), early CD8 effector cluster 16, and GZMB CD8 effector cluster 8 (**Fig 5D**), when compared to the opposing signature. In contrast, the opposing, non-effector phenotype was primarily enriched for transitioning CD8 clusters (6 and 17) and functionally exhausted CD8 cluster 13, with a slight enrichment for GZMK cluster 3. Post-infusion cells sharing TCRs from the precursor effector sorting scheme were also significantly more clonally expanded (Kolmogorov-Smirnov test, p < 0.001; **Fig 5E**). These analyses confirmed that the transcriptional profile associated with precursors of effector lineages is also reflected on the cell surface and demonstrated that the use of surface markers identified by our analyses could be utilized to enrich GMP cells primed to become potent cytotoxic effectors.

After confirming that sorting for this pre-effector signature can enrich for cells with effector GMP product precursor-associated TCR lineages, we assessed whether the pre-effector signature was associated with post-infusion CAR T cell expansion as assessed by qPCR. To determine how a patient’s GMP product aligns with our identified pre-effector signature, we utilized a gene set module score to identify cells in the GMP product with expression profiles most comparable to our TCR-defined precursor lineages. When characterizing GMP infusion products based on the percentage of total T cells or CD8 T cells that matched this expression profile, we found that the percentage of effector precursor CAR T cells of the GMP product correlated positively and significantly with CAR T cell expansion at peak expansion time points specifically as well as at weeks 2 and 3 post infusion (**Fig 5F**). Interestingly, correlations between our transcriptional estimates of effector precursor abundance and expansion were even stronger when excluding patients who exhibited only minimal residual disease prior to infusion (patients 3, 4, 6, and 11; **Fig S5**), highlighting the important relationship between CAR T cell fate and antigen availability.

## Discussion

Our study characterizes and tracks the significant heterogeneity in pre-infusion CAR T cells. Upon infusion into the patient, different CAR T cell subsets lead to divergent differentiation trajectories. The first trajectory involves functional effector differentiation, characterized by expression of conventional cytotoxic genes such as granzymes, *PRF1*, and *NKG7*, from a highly proliferative state of the GMP product. Another group demonstrated a similar differentiation trajectory of proliferation in pre-infusion and early post-infusion CAR T cells leading to potent cytotoxic effector T cell function post infusion in both BCMA- and CD19-CAR T cells (19). As expected, this highly cytotoxic effector state eventually culminated in T cell exhaustion, with the expression of *TOX* and other inhibitory proteins, and cell death as evidenced by an upregulation in *CASP8*. Alternatively, the other trajectory indicates rapid development of these same exhaustion and cell death signatures soon after infusion. By using the endogenous TCR as a method to track CAR T cell lineages, we discovered that the cells of the GMP product giving rise to optimal functional effector phenotypes correspond to a unique subset with a statistically robust transcriptional signature; the presence of this signature in CAR T cells of the GMP product consequently impacts functional effector differentiation and proliferative capacity post infusion. An elegant prior study from Sheih and colleagues used a similar approach to track CD19-CAR T cell lineages in two adult subjects with relapsed and refractory NHL (8). Using paired TCR chain sequences from single cell analysis, they identified CAR T cell clonotypes in the pre-infusion product that either expanded (increased relative frequency, IRF) or declined (decreased relative frequency, DRF) upon infusion. Comparative gene expression analyses between the pre-infusion IRFs and DRFs revealed that the IRF clones displayed higher expression of cytotoxic genes such as *GNLY* and many granzymes, chemokines (*CCL4* and *CCL5*), and the cytokine *IFNG*. Although we present a larger cohort of patients, the substantially greater TCR repertoire diversity characteristic of the pediatric population compared to adults (**Fig S6**) poses significant challenges for identifying shared TCRs between the GMP product and post-infusion CAR T cells. Regardless, the lineages we identified, when analyzed in the context of over 180,000 single-cell expression profiles, allowed us to dissect the specific features of pre-infusion CAR T cells that led to particular functional outcomes post infusion. More broadly, our data demonstrate that despite all CAR T cells of the GMP product having experienced the same product preparation (i.e., activation, viral transduction, *ex vivo* expansion, and cryopreservation) and recognizing the same antigen, they may yet be primed for divergent cell fates.

We were surprised to observe *TIGIT* upregulation in the GMP product precursors of the functional effector population, as this gene is known as a negative regulator of T cell function. TIGIT functions primarily to inhibit T cell effector function by competing with CD266 for binding to CD155 on dendritic cells (20). Adding to its inhibitory potential, TIGIT also prevents homodimerization of CD266 to directly restrain costimulatory signaling (21). The CAR T cells that express TIGIT highly may benefit from its inhibitory effects by limiting the strength of early effector responses that could ultimately lead to a premature dysfunctional state. Under this assumption, overstimulated CAR T cells without TIGIT would become overactivated and, subsequently, exhaust themselves early in the treatment window. The concept of preventing early effector differentiation to delay CAR T cell exhaustion has been suggested previously; for instance, integration of the CAR vector into the *TRAC* locus during product generation abates tonic signaling and impedes the onset of exhaustion (22). These data demonstrate the potential utility in harnessing the role of inhibitory molecules to increase the magnitude and persistence of effector responses.

While our data provide additional context to the transcriptional programs that drive optimal CAR T cell effector function, there are a few limitations within our study. These include foremost sampling issues, since the isolated CAR T cells from post-infusion blood draws only represent a small proportion of the total CAR T cell population, especially at earlier time points when the cells were still expanding. Likewise, sampling only peripheral blood and bone marrow misses some lineages that home to lymphoid organs, effectively reducing the number of lineages available for analysis. Additionally, intensive pre-treatment of the patients participating in the study adds another layer of complexity to the analysis, as these treatments may have altered their baseline T cell states, making it more difficult to elucidate clinical correlates with our GMP precursor phenotype. However, despite this non-exhaustive sampling regime and diverse patient history, we still identified a robust gene signature unique to the pre-effector CAR T cells of the GMP product. Lastly, second generation CAR T cells with a CD28 costimulatory domain have been shown to generate a distinct transcriptional profile in transduced T cells when compared to 4-1BB CAR-transduced cells (23). Thus, it is possible that our identified gene signature is specific for CARs with 4-1BB costimulatory domain, though we would consider this only a minor limitation since 4-1BB CARs are FDA approved and are widely used in preclinical as well as clinical studies.

Our data could have significant implications not only for CD19-CAR but also other CAR T cell-based therapies. The classifier, module score, and enrichment schemes generated based on the gene expression differences between pre-effector CAR T cells and the other GMP cells could potentially be used prior to treatment, after generating the CAR T cell product, to predict CAR T cell product efficacy. In addition, these approaches could assist in deciding if CAR T cell therapy should be combined with other agents or abandoned altogether for particular cases. Moreover, our findings suggest that only a small proportion of CAR T cells within the GMP product determines its efficacy. Using the genes identified in our pre-effector signature, a suite of surface markers could potentially be used to enrich the most efficacious CAR T cells prior to infusion and deplete cells destined for a dysfunctional fate. In conclusion, we provide an innovative method to identify gene signatures for other CAR T cell products by leveraging endogenous TCR sequences within an integrative analytical process (**Fig 6**). Our data provide unique insight into CAR T cell biology in humans and sets the stage to explore opportunities to improve current CAR T cell therapy approaches for a broad range of malignancies.

**Figure 6.**
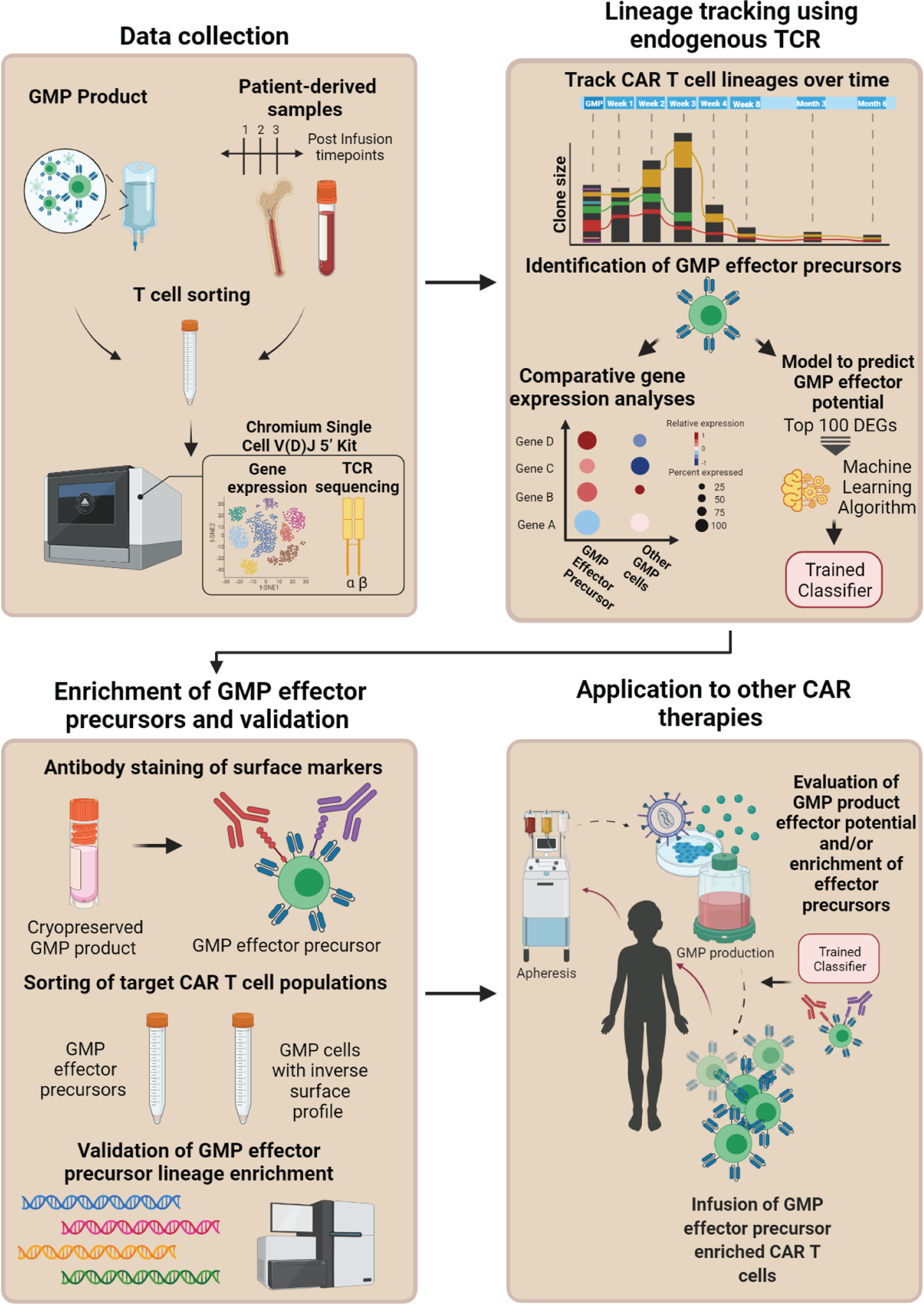
Endogenous TCR tracking as a broadly applicable method for CAR T cells. Schematic overview of experimental approach and analytical pipeline for: 1) identifying signatures associated with precursors of potent cytotoxic effectors in CAR GMP products, 2) evaluating effector potential of CAR GMP products, and 3) enriching CAR GMP products to maximize therapeutic potential.

## Materials and Methods

### Study design and participants

The samples were obtained from subjects enrolled on a single institution Phase I/II clinical study evaluating the safety and efficacy of escalating doses of autologous CD19-CAR T cells in pediatric/adolescent & young adult subjects ≤21 years old with relapsed/refractory CD19-positive B-ALL (SJCAR19; NCT03573700). The protocol was approved by the St. Jude Children’s Research Hospital institutional review board (IRB). Written informed consent/assent were obtained from all participants/parents in accordance with institutional guidelines and the Declaration of Helsinki. The clinical grade lentiviral vector encoding the CD19.4-1BBz CAR, and CD19-CAR T cell products were manufactured at the Children’s Good Manufacturing Practice facility of St. Jude. The study, lentiviral vector, and the manufacturing of autologous CD19-CAR T cells is described in detail elsewhere (11).

Protocol treatment included lymphodepletion (fludarabine [25mg/m^2^, days −4 to −2] and cyclophosphamide [900mg/m^2^, day −2]) followed by CAR T cell infusion (day 0). The first six patients received 1×10^6^ CAR-positive T cells/kg, and starting with patient 7, 3×10^6^ CAR-positive T cells/kg were infused. Response, at 4 weeks post infusion, was categorized as complete response (CR), either minimal-residual disease (MRD)-negative or MRD-positive, or no response (NR). When available for a given patient, MRD-testing included flow-cytometry, RT-PCR and/or next-generation-sequencing (NGS; Adaptive Biotechnologies) techniques.

Peripheral blood samples analyzed for this study were collected at weeks 1-4, 8, and months 3 and 6 post infusion; bone marrow aspirates were obtained at week 4 and month 3 post infusion. qPCR assays for the CD19-CAR transgene were performed as previously described (11).

### Sample processing and T cell sorting for single-cell analysis

PBMCs were separated from whole blood via centrifugation in a BD Vacutainer® CPT™ Mononuclear Cell Preparation Tube (#362761), and remaining red blood cells (RBCs) were lysed for 1 minute at room temperature. Bone marrow cells were collected, and RBCs were lysed for 5 minutes at room temperature. Cryopreserved aliquots of the GMP cell product were thawed at 37° C and washed prior to staining. Cells were blocked with Human TruStain FcX (Biolegend; Cat# 422302) and then stained with a human CD19-CAR Detection reagent (Miltenyi; Cat# 130-115-965) for 10 minutes at room temperature. Cells were then washed twice and incubated with an anti-biotin antibody to label the CD19-CAR detection reagent (Miltenyi; Cat# 130-111-068) and a cocktail of surface antibodies targeting CD45, CD3, CD14, CD16, CD8, and CD4 for 10 minutes at room temperature. Cells were washed twice, stained with DAPI, and then resuspended in fluorescence activated cell sorting (FACS) buffer for cell sorting using the FACS Aria III (BD Biosciences). Total CD3+ T cells were sorted, except when the CAR+:CAR- or CD4:CD8 ratios were highly skewed in GMP product samples. In this case, populations were sorted separately to be combined at equal cell ratios for GMP product samples. For post-infusion samples, only total CD3+ T cells were sorted.

The antibodies used were CD45-FITC (BD Biosciences; Cat# 555482), CD3-APC (Tonbo Biosciences; Cat# 20-0038-1500), CD14-APC-Cy7 (BD Biosciences; Cat# 333945), CD16-APC-Cy7 (BD Biosciences; Cat# 557758), CD8-BV510 (BD Biosciences; Cat# 563919), CD4-BV786 (Biolegend; Cat# 317442).

### Single cell gene expression and V(D)J sequencing

Sorted cells from each sample were counted and assayed for viability via hemocytometer. Because GMP samples were always obtained after cryopreservation, sorted CD4+ and CD8+ populations were sometimes differentially pooled in order to obtain specific CD4:CD8 ratios; however, post-infusion samples were always analyzed fresh, and the CD4:CD8 ratios remained unmanipulated in order to maintain an accurate representation of the CD4+ and CD8+ CAR T cell response dynamics within patients. Sorted cells were processed using the 10X Genomics Chromium controller and the Chromium Single Cell V(D)J 5’ reagents kits (v1) (10X Genomics; Part# 1000014/1000006). T cell receptor V(D)J cDNA was enriched using the Chromium Single Cell V(D)J Enrichment kit for human T cells (10X Genomics; Part# 1000005). V(D)J libraries and 5’ Gene Expression libraries were generated using 10X Genomics library preparation kits. Quantification and quality assessment were completed using the Agilent Tapestation and Agilent High Sensitivity DNA reagents (Agilent; Part# 5067-5593) and Screen Tapes (Agilent; Part# 5067-5592). Libraries were sequenced on the Illumina NovaSeq platform (gene expression sequencing configuration: 26-8-0-91; TCR sequencing configuration: 150-8-0-150).

### Single cell gene expression and V(D)J analysis

Single-cell gene expression data were processed using CellRanger (v.3.1.0, 10X Genomics) with the corresponding GRCh38 reference (v.3.0.0) modified to include the first 825 nucleotide bases of the CD19-CAR transcript. Resulting gene expression matrices were aggregated, again using CellRanger with default parameters (i.e., with depth normalized by the number of mapped reads), resulting in an average of 46,143 reads per cell, over 95% of reads within cells, 1,886 median genes per cell, and 5,955 median UMI counts per cell. Single-cell TCR data were processed using the same version of CellRanger with the corresponding GRCh38 V(D)J reference (v. 3.1.0). Across all V(D)J reactions, the median number of mean TCR read pairs per cell was 11,179 (minimum: 4,419). All included gene expression and V(D)J reactions passed CellRanger quality control metrics.

Aggregated gene expression data were subsequently analyzed using Seurat (24) within the R statistical environment, including cells with a minimum of 300 detected genes and genes found in a minimum of three cells. Standard filtering and processing procedures were utilized prior to downstream analyses. Specifically, we excluded potential doublets and dying cells by filtering out cells with >= 5,000 genes and >= 10% mitochondrial content, respectively. Data were log-normalized using default parameters. We then excluded any cell that did not contain at least one CD19-CAR UMI in order to exclusively analyze transcriptionally defined CD19-CAR T cells. CD19-CAR T cells utilized for downstream analyses exhibited 11,259 median UMI counts per cell, 2,992 median genes per cell, and a median percent of expression owed to mitochondrial genes of 4%.

We identified the top 2,000 variable features using the *vst* method after excluding TCR and IG genes, and data were subsequently scaled using default parameters. In order to correct for potential batch effects accruing over the 1.5-year course of data acquisition from fresh samples, we used the fastMNN algorithm (25), as implemented in Seurat, across each 10X reaction with default parameters. The resulting *mnn* reduction dimensions were subsequently utilized to identify transcriptional clusters via Seurat’s implemented shared nearest-neighbors approach and to generate Uniform Manifold Approximation and Projection (UMAP), again using default parameters.

To identify lineages, we leveraged the TCR sequencing information associated with our single-cell gene expression data. Briefly, we integrated the filtered contig annotations provided by CellRanger with the processed gene expression objects by linking TCR cell barcodes to gene expression cell barcodes. For each patient, we then defined potential clonal lineages using four distinct approaches (visualized in **Fig S3A**). The first two approaches were *α only*, classifying two CAR cells as lineages when all α observed alleles match exactly while disregarding the β chain, and *β only*, classifying two CAR cells as lineages based on matching in only the β chain. Neither of these approaches fully represents the TCR by neglecting the contribution of the other chain. Since cells can differentially express distinct alleles of both the α and β chains of the TCR, the strictest definition of a lineage would be cases where all alphas and all betas must match between two or more cells (scheme 4 in **Fig S3A**). We detected the fewest number of lineages when requiring exact matches of all alleles (**Supplementary Table 3**), and thus defined lineages using a “one-from-each” approach (with one α & one β), where cells that match their most highly expressed (as a stringency filter) α and β chains are designated as lineages.

Pseudotime analysis was conducted using Monocle2 (version 2.20.0) which utilizes DDRTree (Discriminative Dimensionality Reduction via learning a Tree) for dimensionality reduction (26). DDRTree is a reversed graph embedding technique to reduce the data’s dimensionality and make the single cell data into a tree format, so that the trajectories can be visualized and the pseudome calculated. Downsampling was necessary for this analysis due to computational limitations. We randomly selected 368 cells (the number of all the cells at the month 3 time point) from each time point, and we additionally added all 840 cells with TCRs matching known pre- to post-infusion lineages regardless of cluster designation. Pseudotime states were generated based on internal clustering by the pseudotime analysis.

## Statistical analyses

Differential expression analyses were performed using the non-parametric Wilcoxon rank sum test with the Seurat (24) package (version 4.0.1) in the R statistical environment (version 4.1.0) using the *FindMarkers* function (min.pct = 0.2 and logfc.threshold = 0.2). These comparisons included global (each cluster versus all others) and pairwise differential expression analyses across transcriptional clusters for functional annotation, GZMK expression differences between cluster 3 and 8, and comparisons between the transcriptional profile of GMP CD8+ CAR T cells with TCR lineages linked to clusters 3 and 8 versus all other CD8+ GMP CAR+ cells. We corrected the p-values for multiple testing with the Bonferroni procedure, as suggested by Seurat documentation, and considered genes with an adjusted p-value < 0.05 as statistically significant.

The Kolmogorov–Smirnov test was used to evaluate differences in clone sizes across comparator groups (e.g., **Fig S3B**, **Fig 5E**). To construct a classifier for predicting CAR T cell effectors of GMP cell products, the top 100 differentially expressed genes between downsampled pre-effector and the other downsampled CD8+ T cells of GMP products were selected for training. A support vector machine (27) with a radial kernel was used for the classification. We utilized 1,000 iterations to obtain a robust result, and the LOOCV (Leave One Out Cross Validation) approach was used in each iteration.

As another, broader approach to identify cells of the CAR product that may have been effector precursors in the absence of TCR lineage information, we defined a gene set module using the top 20 upregulated genes in the CD8+ CAR T cells of the GMP product with lineages ending in the functional effector CD8 T cell clusters 3 and 8 compared to all other GMP CD8+ CAR+ cells. The module score was calculated for each cell of the GMP product using the *AddModuleScore* function in Seurat with default parameters. Upon investigating the distribution of module scores across our data, we selected a threshold of >=0.4 to identify putative precursor effectors from outside of our lineage analyses. We then calculated the percentage of CD8+ cells within the GMP product that met this threshold for each patient. Spearman’s rank correlations between these percentages and qPCR-determined CAR abundance were calculated using the *corr.test* function (use = “complete.obs”) from the R *psych* package (28). P-values were adjusted for multiple testing using the FDR approach (29).

The comparison of TCR diversity between our cohort and a previously published study on adult Non-Hodgkin’s lymphoma (8) was done based on TCRβ chains. The TCR data obtained via 10X Genomics were accessed from the NCBI GEO database (Accession: GSE125881) and compared to the TCRβ chains obtained via 10X Genomics for this cohort using the Shannon-Wiener index. A Wilcoxon test was performed to assess statistical significance of the difference.

### Effector GMP product precursor signature validation

Cryopreserved GMP product cells were thawed at 37°C and washed. Cells were, then, stained with the CD19-CAR Detection reagent (Miltenyi; Cat# 130-115-965) for 10 minutes at room temperature followed by staining with an anti-biotin antibody conjugated to APC (Miltenyi; Cat# 130-110-952) and a cocktail of antibodies (CD3, CD8, CD27, CD62L, CD25, and TIGIT) and a viability dye (Tonbo Biosciences; Cat# 13-0870-T100). After staining for 10 minutes at room temperature, the cells were washed twice and resuspended in FACS buffer for sorting on the FACS Aria Fusion (BD Biosciences). CD8+ CAR T cells with either the predicted effector surface profile (TIGIT+, CD62L^lo^ and CD27-) or the opposite, non-effector precursor profile (TIGIT-, CD62L+, and CD27+) were sorted into complete RPMI media. Cells were lysed with Trizol for bulk TCR repertoire sequencing.

Antibodies used were anti-human CD3-APC-H7 (BD Pharmingen; Cat #560176), anti-human CD8-BV785 (Biolegend; Cat # 344740), anti-human CD27-PE-CF594 (BD Horizon; Cat # 562297), anti-human CD62L-BV421 (Biolegend; Cat # 304828), anti-human CD25-VioBright FITC (Miltenyi Biotec; Cat# 130-113-283), and anti-human TIGIT-PE (Biolegend; Cat# 372703).

### Bulk TCR sequencing and analysis

Bulk repertoire sequencing was performed using the 5’RACE protocol adapted from Egorov and colleagues (30). In brief, total RNA was isolated from sorted cells with Trizol reagent (Invitrogen; Cat# 15596026) using the manufacturer’s protocol. cDNA synthesis was performed with SmartScribe kit (TakaraBio; Cat# 639537) with C-segment specific primers and template switching oligonucleotide with randomized UMI sequence. cDNA synthesis product was purified using Ampure XP kit (Beckman Coulter; Cat# A63880), and amplified in two rounds of PCR with Q5 HotStart high-fidelity polymerase kit (NEB; Cat# M0493S). Adapters for Illumina sequencing were ligated with KAPA HyperPrep kit (Roche; Cat# 07962363001). The libraries were sequenced on the Illumina NovaSeq platform (2×150 paired end sequencing).

Sample demultiplexing and UMI-guided assembly were conducted using MiGEC (v.1.2.9) *CheckoutBatch* (with -ute flags) and *AssembleBatch* (with --force-overseq set to 1) functions, respectively (31). Individual sample assemblies were further parsed and annotated using the MiXCR (v.3.0.13) *analyze amplicon* function (--species hs --starting-material rna --5-end no-v-primers --3-end c-primers --adapters adapters-present --receptor-type tcr) (32). Resulting TCR outputs were then converted to VDJtools format using the VDJtools (v.1.2.1) software suite (33) for downstream analysis. To characterize the bulk TCR repertoires from the signature sort experiments in the context of the post-infusion single-cell TCR data, we identified exact nucleotide matches in the CDR3 regions between bulk β chain sequences and single-cell β chain sequences, using the most highly expressed β chain in cases where multiple betas were identified in an individual cell. After identifying exact β chain matches between the bulk GMP data and post-infusion single-cell data, we used those matches to infer the future transcriptional clusters of the β lineages for each sample, which we represented in terms of proportions of the bulk β sequences with exact matches (**Fig S4B**). We then calculated log_2_ fold changes between the two sort schemes (**Fig 5D**). The Kolmogorov–Smirnov test was used to evaluate differences in clone sizes across the two sort schemes (**Fig 5E**).

### Data and code availability

Single-cell gene expression data, single-cell TCR data, and bulk TCR data will be made publicly available prior to peer-reviewed publication. Relevant clinical data are included in the manuscript or referenced elsewhere (see **Supplementary Clinical Information**). qPCR data for assessing CAR expansion are included as Supplementary Table 2.

## Data Availability

Single-cell gene expression data, single-cell TCR data, and bulk TCR data will be made publicly available prior to peer-reviewed publication. Relevant clinical data are included in the manuscript or referenced elsewhere (see Supplementary Clinical Information). qPCR data for assessing CAR expansion are included as Supplementary Table 2.

## Acknowledgements

This work was supported by the National Institutes of Health (NIH)/National Cancer Institute grant P30CA021765, NIH grants U01AI150747 and R01AI136514 (PGT), the American Society of Transplantation and Cellular Therapy (AT), the American Society of Hematology (AT), the Key for a Cure Foundation (PGT), the Mark Foundation ASPIRE Award (PGT), and the American Lebanese Syrian Associated Charities (SG, PGT). Part of the laboratory studies were performed by the Center for Translational Immunology and Immunotherapy (CeTI^2^), which is supported by SJCRH. The content is solely the responsibility of the authors and does not necessarily represent the official views of the NIH. We thank the referring physicians, the staff of the clinical research office for assisting with conducting the clinical study, the staff of the Human Applications Laboratory and GMP facility for assisting in CAR T-cell production and analysis, and the staff of the Department of Bone Marrow Transplantation and Cellular Therapy for their excellent patient care. We thank David Cullins and Sagar Patil for assistance with flow and sorting, Sarah Schell and Carrie Henson for performing the qPCR assays, MaCal Tuggle-Brown for assistance with sample processing, and Mireya Paulina Velasquez and Swati Naik for helpful discussions and suggestions. We also thank the patients who participated in this study and their caregivers, who entrusted the care of their children to us. Graphical schematics were created using Biorender.com, for which we have a license.

---

**Figure S1.**
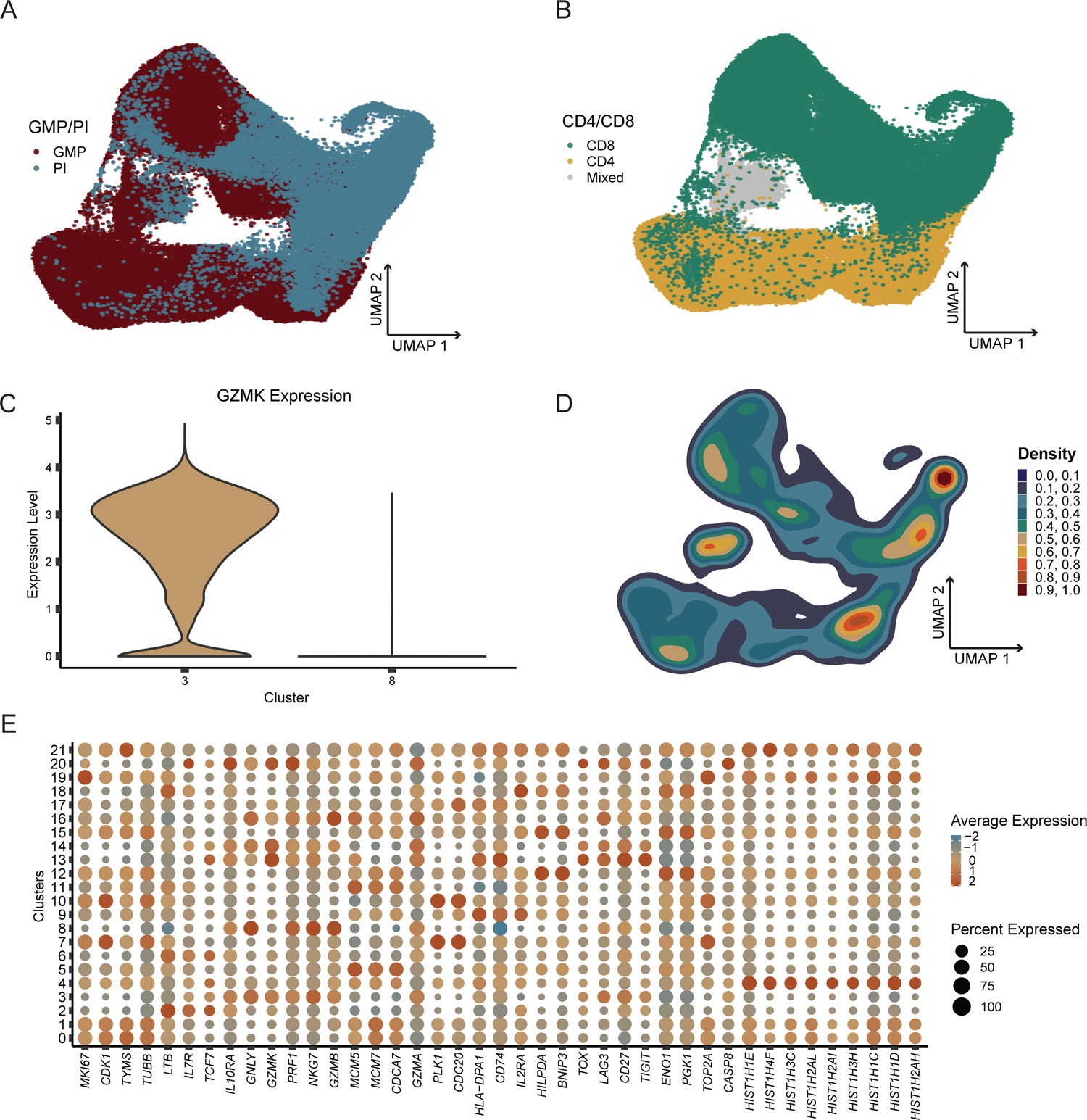
Single cell transcriptomics of pre-and post-infusion CAR T cells. **A-B**, UMAP plots of pre- and post-infusion CAR T cells colored by GMP and post-infusion status (A) or CD4/CD8 status (B). **C**, Violin plot comparing expression of GZMK between transcriptional clusters 3 and 8. **D**, UMAP plot colored by cellular density. **E**, Dot plot visualizing the relative expression of key markers utilized in annotation of transcriptional clusters (see Fig 1C). Dot size corresponds to percent of cells expressing a gene.

**Figure S2.**
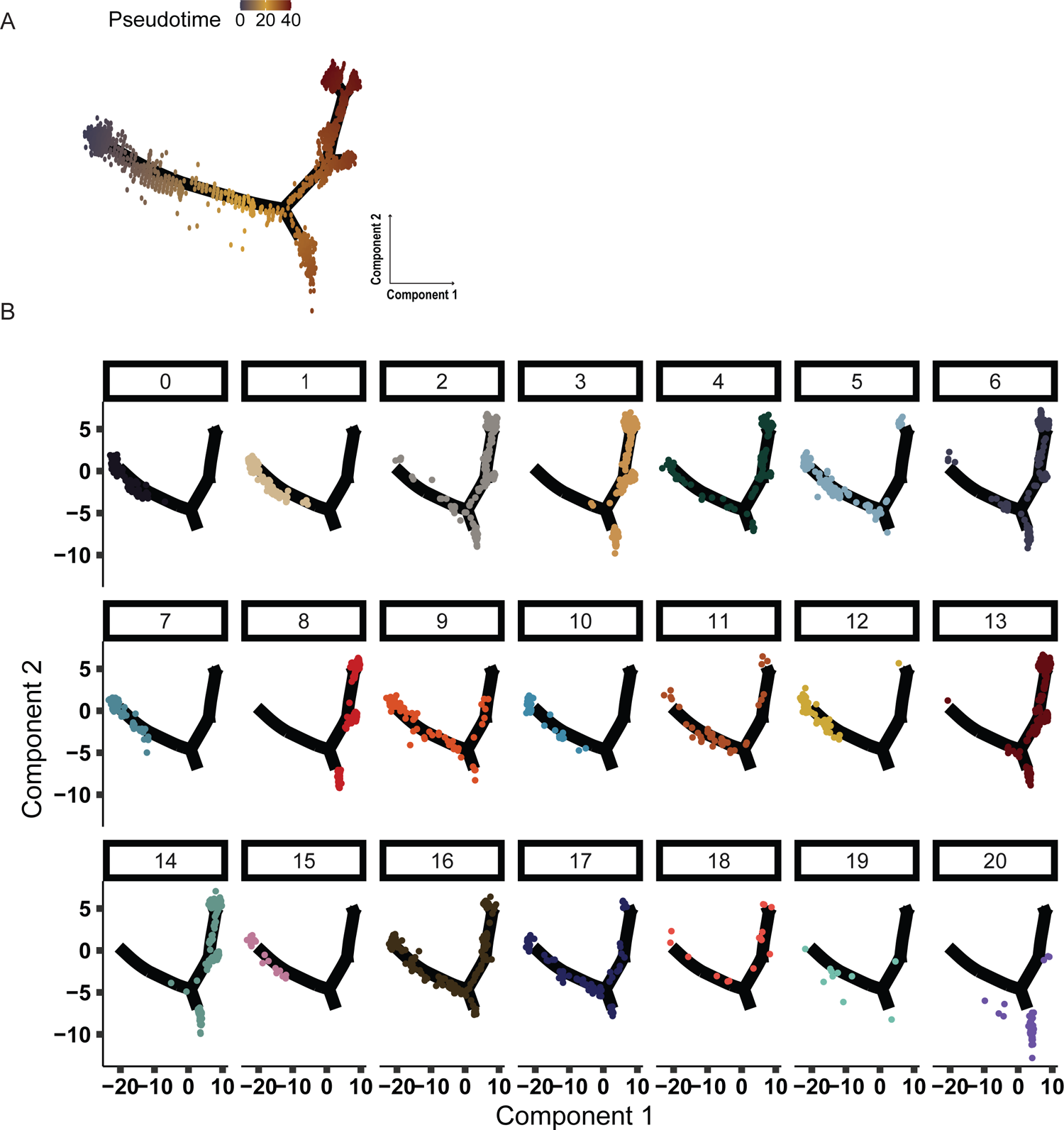
Characterization of CAR T cells in pseudotime. **A**, Monocle pseudotime map of CAR T cell transcriptional trajectory, colored by pseudotime gradient. **B**, Monocle pseudotime maps depicting each transcriptional cluster.

**Figure S3.**
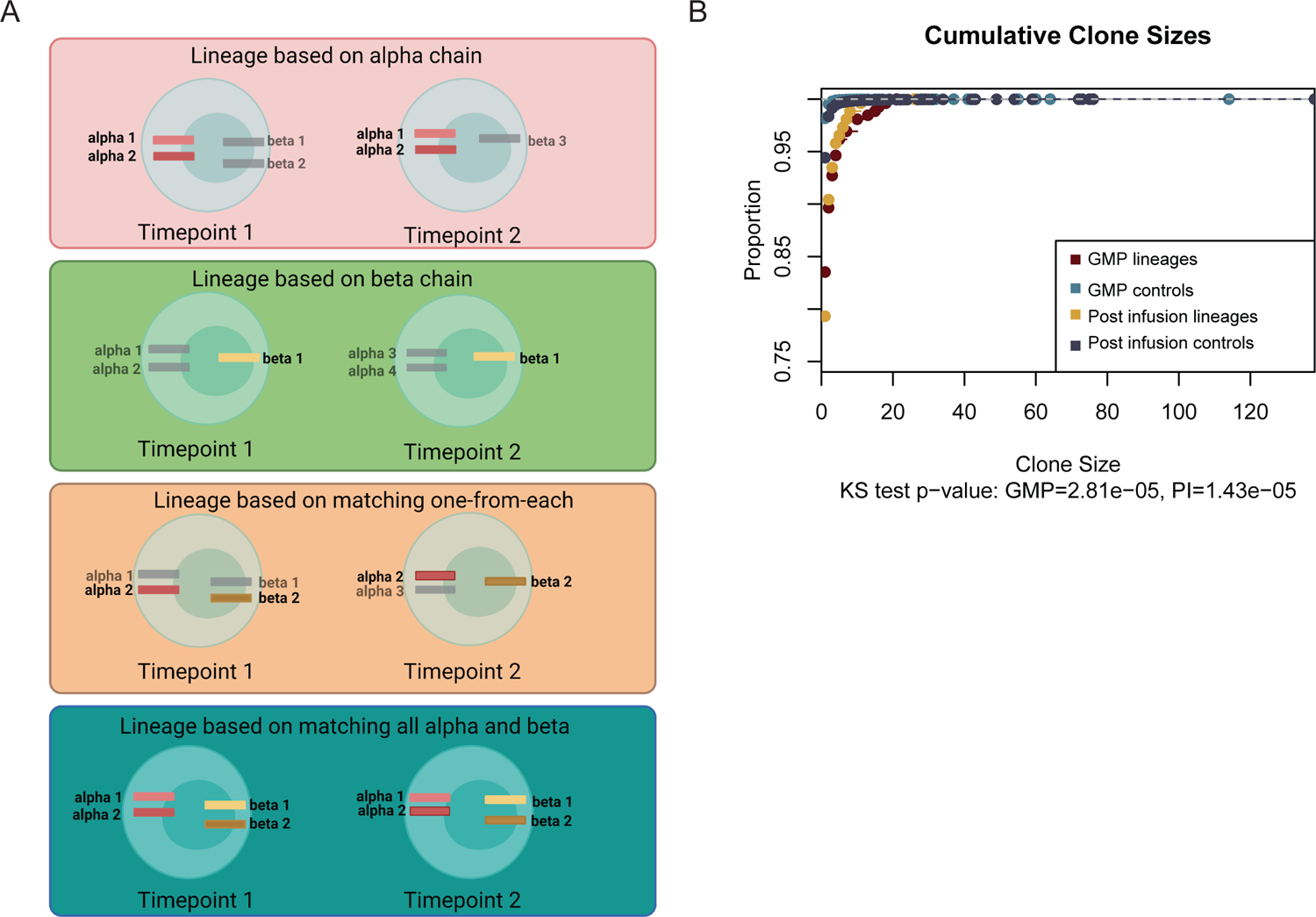
TCR lineage tracking. **A**, Different TCR α and β matching schemes. The complementarity determining region 3 (CDR3) of each α and β chain is sequenced to determine expressed alleles of each chain. Scheme 1 (red) describes classifying two CAR cells as in the same lineage when all observed α alleles match exactly while disregarding the β chain. Scheme 2 (green) assigns cells to lineages based on complete matching in only the β chain. Neither scheme 1 nor 2 fully represents the TCR by neglecting the contribution of the other chain. Since cells can differentially express distinct alleles of both the α and β chains of the TCR, the strictest definition of a lineage would be cases where all alphas and all betas must match between two or more cells (scheme 4; bottom). We detected the fewest number of lineages when requiring exact matches of all alleles (Supplementary Table 1). Thus, we defined lineages using a “one-from-each” approach (scheme 3; orange), where cells that match their most highly expressed (as a stringency filter) α and β chains are designated as lineages. **B**, Cumulative clone sizes of: GMP (dark red) and post-infusion (yellow) CAR T cell lineages, and GMP (light blue) and post-infusion (dark blue) CAR T cells without observed lineages (controls). Graph represents the proportion of cells (y-axis) cumulatively encompassed by increasing clone sizes.

**Figure S4.**
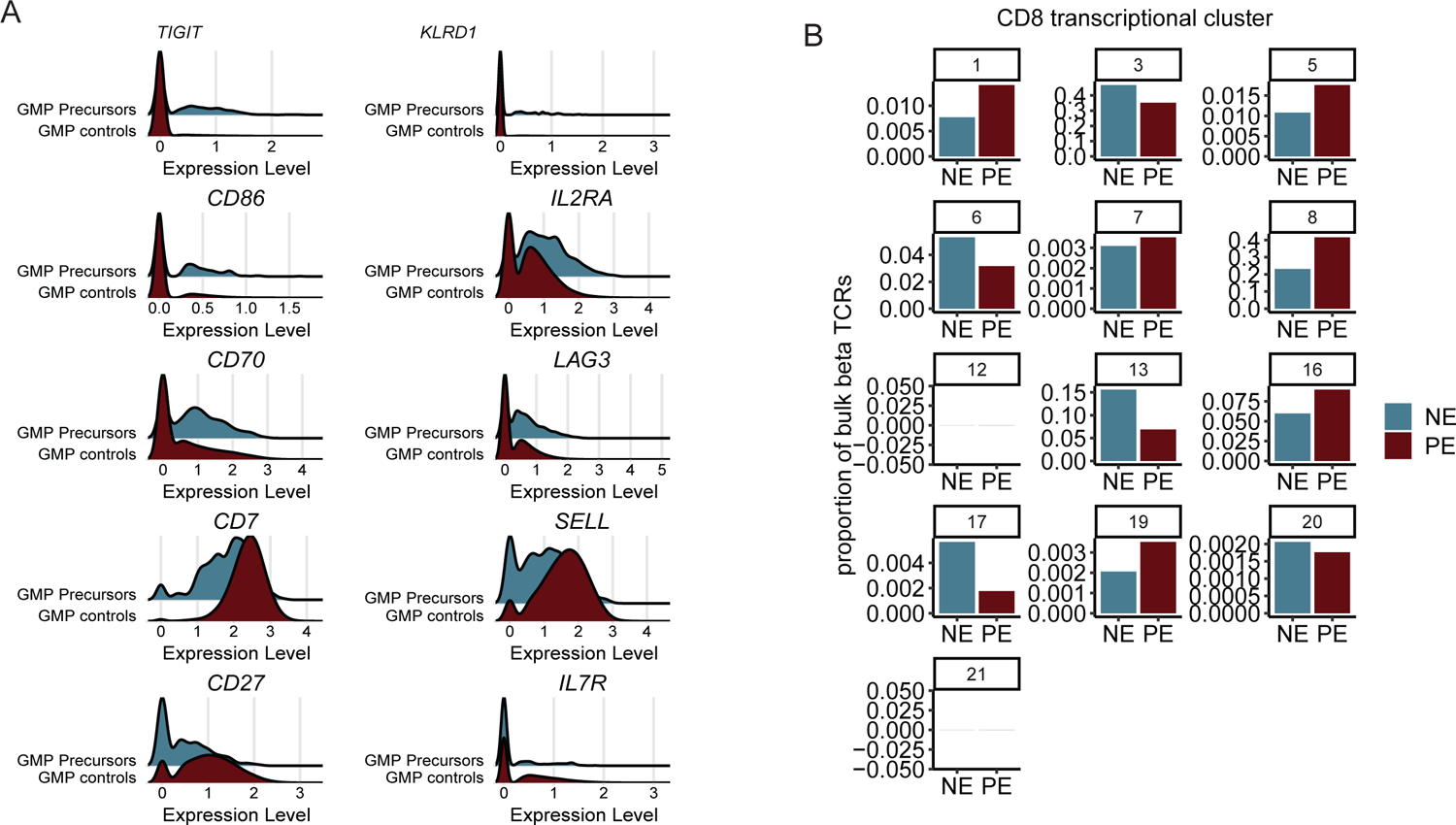
Selection of surface targets for pre-effector CAR T cell surface phenotype validation and results of enrichment. **A**, Histograms comparing expression of a subset of genes encoding surface proteins in effector precursors (red) and non-effector precursors (green). **B**, Bar plots representing the inferred proportions of post-infusion CD8 transcriptional clusters linked to Patient 11 GMP CAR T cells sorted to approximate either the pre-effector signature (PE) or the opposing, non-effector signature (NE). TCRβ chains were sequenced in bulk from each sort, and cluster-annotated post-infusion cells from CD8 clusters that shared those β sequences were used to infer the post-infusion transcriptional cluster those lineages would track to.

**Figure S5.**
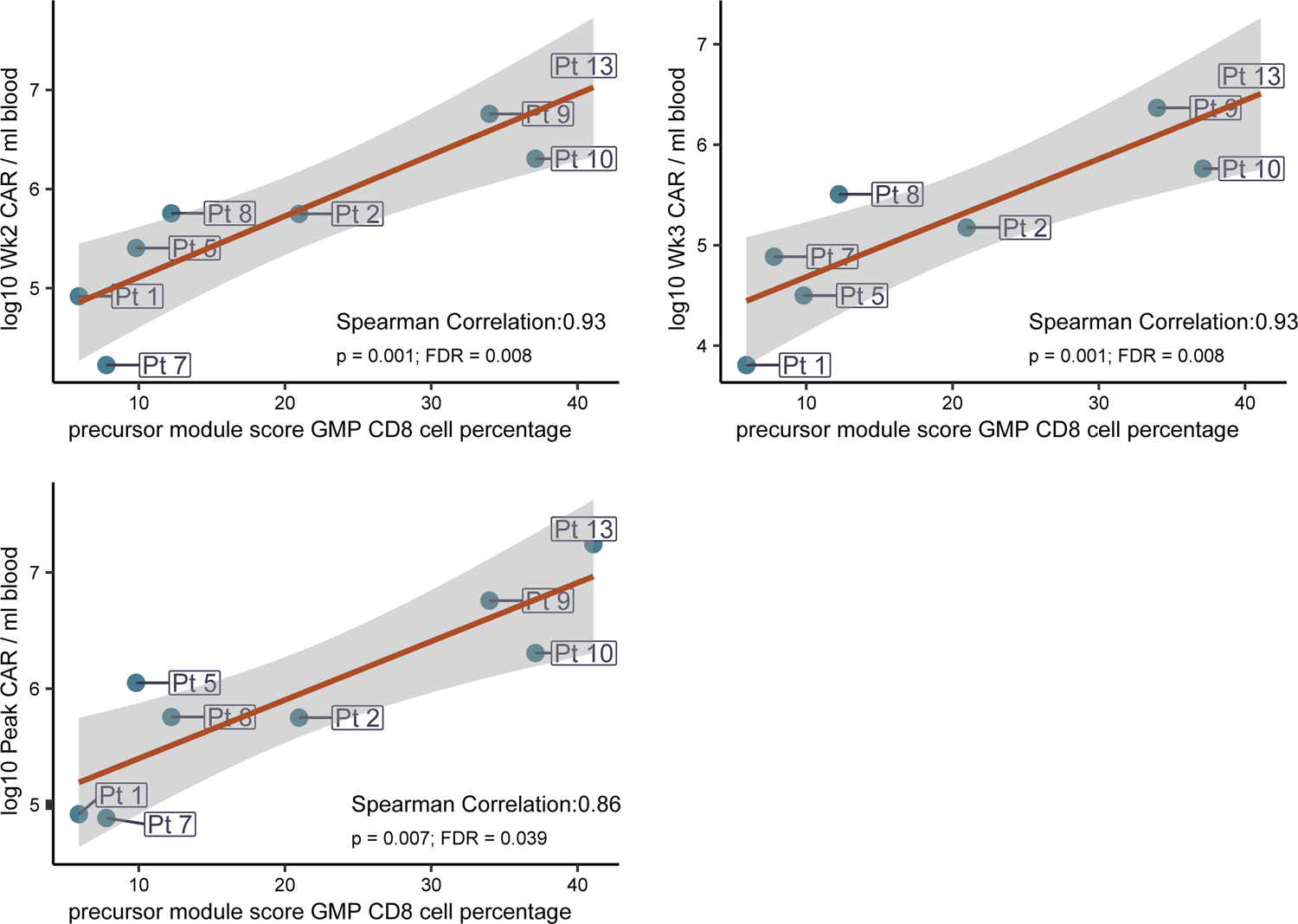
Inferred precursor effector abundance of GMP products correlates strongly with CAR T cell expansion when excluding patients with only minimal residual disease. Correlation plots comparing the percentage of inferred effector precursors in the GMP product to CAR abundance as assayed by qPCR, and represented in terms of CAR copies per ml of blood, at week 3, week 2, and peak expansion time points. These are replicates of the correlations presented in Figure 5F, but patients with only minimal residual disease prior to infusion were excluded from these correlations.

**Figure S6.**
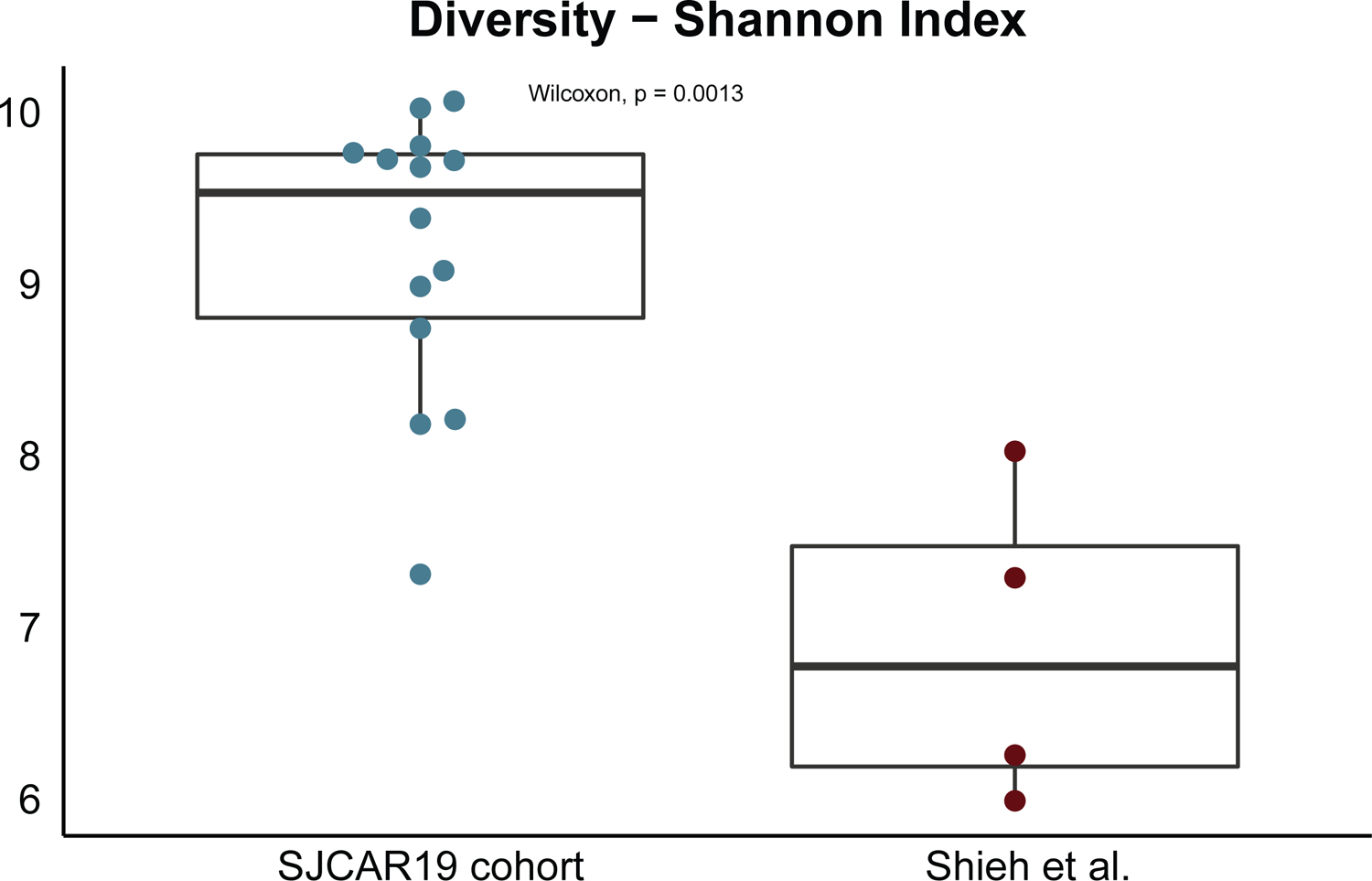
GMP products generated from pediatric patients exhibit extreme clonal diversity compared to those generated from adult patients. Box plots comparing the β chain diversity of CAR T cell TCRs from the GMP product in pediatric patients from the current study compared to those from adult patients in a previously published study (Sheih et al 2020, Nature Communications). The Shannon-Wiener index was calculated for each patient, and a Wilcoxon test was performed to evaluate statistical significance.

## Supplementary clinical information on patients enrolled on SJCAR19

(A Phase I/II Study evaluating CD19-specific engineered autologous T cells in pediatric and young adult patients ≤ 21 years of age with relapsed or refractory CD19+ acute lymphoblastic leukemia; NCT03573700) The clinical results of patients 1 to 12 have been reported in abstract form (*Talleur et al; Blood* (*2020*) *136 (Supplement 1): 39–40;* https://doi.org/10.1182/blood-2020-138528) and/or are submitted for publication (Talleur *et al*; manuscript in revision). In addition, the clinical course of patients 1 and 9 have been published (*Himes et al; Br J Haematol. 2021 Aug;194*(*4*)*:701-707.* https://onlinelibrary.wiley.com/doi/full/10.1111/bjh.17662). For patient 0, a GMP CD19-CAR T cell product was manufactured, but the patient did not proceed to the treatment portion of the clinical study due to poor clinical status in the setting of rapidly progressive B-ALL. Patients 13 – 15 were enrolled on the Phase II portion of the clinical trial and received protocol prescribed treatment with lymphodepleting chemotherapy (fludarabine/ cyclophosphamide) followed by infusion of CD19-CAR T cells (3×10^6 CAR+ T cells/kg). These patients were between 5 and 10 years old at time of infusion. Indication for treatment included relapse after hematopoietic cell transplant (HCT), relapse 2 and refractory disease in relapse 1. Pre-treatment disease burden in the marrow ranged from 0-51% blasts by morphology (Patient 13: 32.34%, Patient 14: 27.88%, Patient 15: 0.023% by flow cytometry); none had detectable leukemia in the cerebrospinal fluid. Patient 13 had grade 2 cytokine release syndrome (CRS) post infusion, patient 14 had fever <24 hours from infusion (infusion related reaction) and patient 15 had grade 1 CRS; no one had immune effector cell-associated neurotoxicity syndrome (ICANS). All three patients achieved a complete response (CR) at 4 weeks post infusion, of which two were MRD (minimal residual disease) negative and one MRD positive, respectively. Patients 13 and 14 then proceeded to consolidative allogeneic HCT and both remain alive and in CR at time of last follow-up. Patient 15 experienced leukemic recurrence (CD19-antigen loss) approximately 2-months after CAR T cell infusion, and died secondary to leukemia.

## Notes

### Competing Interest Statement

TLW, HK, SG, JCC, and PGT have patent applications in the fields of T cell or gene therapy for cancer.

### Author Declarations

The samples were obtained from subjects enrolled on a single institution Phase I/II clinical study evaluating the safety and efficacy of escalating doses of autologous CD19-CAR T cells in pediatric/adolescent & young adult subjects <=21 years old with relapsed/refractory CD19-positive B-ALL (SJCAR19, NCT03573700). The protocol was approved by the St. Jude Children's Research Hospital institutional review board (IRB). Written informed consent/assent were obtained from all participants/parents in accordance with institutional guidelines and the Declaration of Helsinki.

