## Supplemental tables 1-4 for "Common trajectories of highly effective CD19-specific CAR T cells identified by endogenous T cell receptor lineages"

**Supplementary Table 1.** Numbers of CD19 CAR T cells analyzed via single-cell transcriptomics per patient and sample.

| Patient | Pre-infusion | Post infusion | | | | | | |
|---|---|---|---|---|---|---|---|---|
| | GMP Product | Week 1 | Week 2 | Week 3 | Week 4 | Week 8 | Month 3 | Month 6 |
| **0** | 1,341 | | | | | | | |
| **1** | 1,885 | | | | | | | |
| **2** | 8,568 | 261 | 830 | 84 | 61 | | | |
| **3** | 8,222 | 169 | 146 | 1 | 4 | | | |
| **4** | 12,280 | 340 | 1,220 | 459 | 434 | 5 | | |
| **5** | 11,988 | 9,510 | 1,027 | 672 | 321 | 114 | 355 | 7 |
| **6** | 8,461 | 144 | 1,444 | 556 | 279 | 5 | 13 | |
| **7** | 7,607 | 36 | 39 | 26 | 38 | 11 | | |
| **8** | 14,840 | 971 | 1,069 | 1,273 | 2,165 | 251 | | |
| **9** | 11,221 | 136 | 2,974 | 874 | 184 | | | |
| **10** | 10,960 | 1,568 | 4,389 | 1,705 | 851 | 1,149 | | |
| **11** | 12,218 | 2,427 | 1,940 | 5,234 | 1,146 | | | |
| **12** | 4,339 | 190 | | | | | | |
| **13** | 4,819 | | | 3,473 | | | | |
| **14** | | 2,855 | 386 | 2,372 | 21 | | | |
| **15** | | 2,478 | 2,148 | 3,143 | 58 | | | |

Black colored boxes indicate samples were not available for analysis. Grey indicates patients that were excluded from GMP-post infusion lineage tracing analysis due to data limitations.

**Supplementary Table 2.** CD19 CAR/ml of blood by week post infusion, as assessed by qPCR.

| Patient | Week 1 | Week 2 | Week 3 | Week 4 | Week 8 | Month 3 | Month 6 |
|---|---|---|---|---|---|---|---|
| 1 | 9,341 | **82,939** | 6,414 | 6,677 | | | |
| 2 | 296,787 | **561,988** | 149,865 | 51,570 | 1,374 | | |
| 3 | **947,250** | 254,181 | 16,461 | 958 | 1,003 | | |
| 4 | 24,967 | **2,165,026** | 421,573 | 121,406 | 3,801 | 4,209 | |
| 5 | **1,122,227** | 254,016 | 31,556 | 36,746 | 114,889 | 174,431 | 0 |
| 6 | 50,700 | **3,369,685** | 894,403 | 376,926 | 17,141 | 2,970 | 2,905 |
| 7 | 7,561 | 16,755 | **76,891** | 8,432 | 872 | | |
| 8 | 314,646 | **569,687** | 321,331 | 91,583 | 89,693 | | |
| 9 | 268,449 | **5,732,386** | 2,328,662 | 339,582 | | | |
| 10 | 1,960,334 | **2,024,664** | 578,331 | 27,864 | 139,449 | | |
| 11 | 236,747 | **2,825,332** | 1,621,885 | 168,308 | | | |
| 12 | 66,126 | | | | | | |
| 13 | 49,091 | **17,544,341** | 4,843,874 | 1,740,770 | | | |
| 14 | **1,640,116** | 69,673 | 471,052 | 2,238 | | | |
| 15 | 395,350 | **520,669** | 81,407 | 7,298 | 10,025 | | |

Grey colored boxes indicate time points with unavailable data. Bolded numbers correspond to peak expansion values for each patient.

**Supplementary Table 3.** Assessing performance of different strategies to define CAR T cell lineages by tracking T cell receptors across time points.

| Patient | α only | β only | one α & one β | Strict |
|---|---|---|---|---|
| 0 | N/A | N/A | N/A | N/A |
| 1 | N/A | N/A | N/A | N/A |
| 2 | 42 | 13 | 9 | 7 |
| 3 | 4 | 1 | 0 | 1 |
| 4 | 119 | 87 | 74 | 65 |
| 5 | 271 | 75 | 32 | 27 |
| 6 | 133 | 96 | 100 | 81 |
| 7 | 13 | 13 | 11 | 11 |
| 8 | 367 | 114 | 70 | 62 |
| 9 | 228 | 52 | 41 | 29 |
| 10 | 566 | 242 | 163 | 139 |
| 11 | 658 | 452 | 385 | 361 |
| 12 | 1 | 0 | 0 | 0 |
| 13 | 72 | 12 | 1 | 1 |
| 14 | 22 | 13 | 11 | 11 |
| 15 | 198 | 175 | 165 | 163 |
| **Total:** | **2,694** | **1,345** | **1,062** | **958** |

The complementarity determining region 3 (CDR3) of each α and β chain were sequenced to determine expressed alleles of each chain. α only: classifying two CAR cells as lineages when all α observed alleles match exactly while disregarding the β chain. β only: classifying two CAR cells as lineages based on matching in only the β chain. Neither of these approaches fully represents the TCR by neglecting the contribution of the other chain. Since cells can differentially express distinct alleles of both the α and β chains of the TCR, the strictest definition of a lineage would be cases where all alphas and all betas must match between two or more cells (Strict). We detected the fewest number of lineages when requiring exact matches of all alleles, and thus defined lineages using a "one-from-each" approach (one α & one β), where cells that match their most highly expressed α and β chains (as a stringency filter) are designated as lineages. N/A= not applicable due to unavailability of either GMP or post-infusion samples.

**Supplementary Table 4.** Assessing clonal expansion among post-infusion CAR T cells. For each patient's post-infusion samples, we show the number of unique clones across a range of clone sizes. E.g., clone size = 1 corresponds to the number of unique clones only found in a single cell.

| Patient | clone size = 1 | 1 < clone size <= 10 | clone size > 10 |
|---|---|---|---|
| 0 | NA | NA | NA |
| 1 | NA | NA | NA |
| 2 | 979 | 11 | 0 |
| 3 | 169 | 0 | 0 |
| 4 | 1,150 | 133 | 1 |
| 5 | 5,184 | 178 | 0 |
| 6 | 1,302 | 185 | 2 |
| 7 | 87 | 4 | 0 |
| 8 | 4,958 | 129 | 1 |
| 9 | 3,323 | 107 | 0 |
| 10 | 7,085 | 396 | 12 |
| 11 | 8,028 | 639 | 4 |
| 12 | 180 | 1 | 0 |
| 13 | 2,824 | 209 | 0 |
| 14 | 4,807 | 156 | 4 |
| 15 | 4,905 | 472 | 43 |
| **Total** | 44,981 | 2,620 | 67 |

N/A= not applicable due to unavailability of post-infusion samples.
